# Surfacing Suicidal Risk Through Simulated Social Interaction: Per-Person Language Model Agents as Communicative Stress Tests

**DOI:** 10.64898/2026.06.04.26354928

**Authors:** Wenpei Shao, Brooke Ammerman, Ross Jacobucci

## Abstract

Suicidal risk may be encoded in everyday communication patterns but diluted in routine digital interactions. We introduce a method for surfacing this latent signal: training per-person language model agents on individuals’ authored text (the on-screen text each participant typed, captured whenever a keyboard was visible in screenshots) and placing those agents in simulated social interactions—a “communicative stress test.” Using data from 79 adults with recent suicidal ideation, we fine-tuned individual LoRA adapters on Qwen3-8B using each participant’s authored text, then placed agents in standardized conversations with probe personas. Agent-generated risk language was associated with EMA-measured suicidal ideation (*r* = .576, *p <* .001), with a single neutral small-talk probe performing nearly as well (*r* = .551). A shuffle control confirmed the signal is person-specific (*r* = .071 when adapters were mismatched), and automated descriptions of participants’ general smartphone activity produced no signal, confirming specificity to interpersonal communication. A prompt ablation demonstrated partial robustness to removal of disclosure-encouraging language (*r* = .430). This proof-of-concept demonstrates that simulated social interaction can amplify latent vulnerability signals, bridging digital phenotyping, generative AI, and suicide theory.

## 1. Introduction

Suicide remains a leading cause of death globally, and predicting who is at risk continues to be one of the most difficult problems in clinical science (Franklin et al., 2017). Despite decades of research, clinicians perform only marginally better than chance at predicting suicidal behavior (Large et al., 2016), and traditional risk factor approaches have reached apparent ceiling effects in predictive accuracy (Belsher et al., 2019). A central challenge is that suicidal ideation fluctuates on the order of hours (Kleiman et al., 2017), and the interpersonal processes theorized to drive it—including perceived burdensomeness and thwarted belonging—are inherently relational, manifesting most clearly in social interaction rather than in static self-report.

Two converging developments create new opportunities for studying these dynamics. First, digital phenotyping (Insel, 2017) has produced rich longitudinal records of how individuals communicate in daily life. Screenomics, which captures smartphone screenshots at five-second intervals (Reeves et al., 2021), provides near-complete records of messages sent and received across all messaging surfaces (e.g., Snapchat, Discord, Messenger, WhatsApp). When combined with ecological momentary assessment (EMA) of suicidal ideation, these data *offer* the potential to trace moment-to-moment relationships between digital communication and suicide risk. In adolescent populations, intensive longitudinal smartphone sensing has shown particular promise: Auerbach et al. (2023) demonstrated that mood disturbances measured via smartphone predicted clinically significant suicidal ideation on a weekly basis, and subsequent work used GPS data to detect next-week suicide risk (Auerbach et al., 2025). However, analyzing raw message content or sensor data for suicide risk faces fundamental limitations: most messages are logistical and routine, and risk-relevant content is sparse and diluted across thousands of everyday communications.

Second, the emergence of large language models (LLMs) capable of producing human-like text has enabled a new class of research: generative agent simulation. Park et al. (2023) demonstrated that LLM-powered agents placed in a shared environment exhibit emergent social behaviors without explicit programming. Subsequent work has scaled this to 1,000 agents parameterized from real interview data, achieving 85% behavioral fidelity (Park et al., 2026). More broadly, LLMs have been used as proxies for human participants—Argyle et al. (2023) introduced “silicon sampling” to simulate survey responses, Aher et al. (2023) replicated classic psychology experiments, and Peters and Matz (2024) showed that LLMs can infer Big Five traits from social media posts (*r* = 0.29; see also Brickman and Oltmanns 2025). Applications to mental health have begun to appear, including agent-based simulations of student wellbeing (Sommuang et al., 2025), frameworks for modeling socio-environmental determinants (Kambeitz and Meyer-Lindenberg, 2025), and simulations of vulnerable users interacting with AI chatbots (Qiu et al., 2025). Louie et al. (2024) validated the use of expert-designed probe personas for patient simulation. Concurrently, there has been a rapid expansion of LLMs applied directly to suicide prevention (Holmes et al., 2025). However, Larooij and Törnberg (2026) offered a critical review arguing that LLMs may exacerbate rather than solve the validation challenges inherent in agent-based social simulation.

A substantial psycholinguistic literature has established that language carries reliable markers of suicidal risk. Seminal work by Stirman and Pennebaker (2001) demonstrated that suicidal poets used more first-person singular pronouns and fewer first-person plural pronouns than non-suicidal poets. Al-Mosaiwi and Johnstone (2018) showed that absolutist thinking—operationalized through words like “nothing,” “completely,” and “always”—is a cognitive-linguistic marker specific to suicidal ideation beyond what negative emotion words alone capture. More broadly, a systematic review (Homan et al., 2022) identified consistent markers across studies, including elevated self-referential language, death-related vocabulary, and reduced future-oriented words. This linguistic signature has been validated at scale in user-generated content on social media: De Choudhury et al. (2016) showed that linguistic shifts in mental-health-related Reddit posts predicted transitions to suicidal ideation, and Coppersmith et al. (2018) developed NLP-based screening for suicide risk from Twitter and Reddit, demonstrating that the same psycholinguistic markers identified in clinical samples generalize to spontaneous online expression. Dictionary-based approaches have been central to translating these findings into applied risk detection, with the Linguistic Inquiry and Word Count (LlVC; Pennebaker et al. 2015) program applied to suicide notes (Pestian et al., 2012), crisis hotline transcripts (Huang et al., 2024), and clinical interviews (Tausczik and Pennebaker, 2010). In clinical messaging contexts, Swaminathan et al. (2023) demonstrated that a two-stage NLP system combining keyword filtering with machine learning reduced crisis message triage time from 9 hours to 8-13 minutes. Prior dictionary-based work (Ammerman et al., 2025) adapted the Swaminathan crisis-language dictionary (Swaminathan et al., 2023) into seven theme-specific sub-dictionaries (suicidal thoughts, non-substance methods, substances, sleep, hopelessness, general risk, and help-seeking) and found that several categories—most consistently general risk and hopelessness—tracked concurrent EMA-measured suicidal ideation and planning at both between- and within-person levels.

The present study bridges digital phenotyping and generative agent simulation by training per-person language model agents on individuals’ authored text and placing those agents in simulated social interactions to surface latent communication patterns associated with suicidal risk (Figure 1). For each participant, we fine-tune a low-rank adaptation (LoRA) adapter on an open-source language model (Qwen3-8B) using their real messages, creating an agent that encodes their communicative style. We then place each agent in standardized conversations with probe personas designed to elicit specific interpersonal dynamics, and score the generated language for risk-relevant content using the dictionary-based approach described above. We conceptualize this as a *communicative stress test* —analogous to cardiac stress testing, where a resting ECG may appear normal but underlying vulnerability becomes visible under exertion. In our paradigm, routine authored text is the resting state, and the simulated conversation is the treadmill. This approach shares a conceptual affinity with recent work using multi-agent AI systems to systematically probe wearable sensor data for clinically relevant biomarkers (Kim et al., 2026), though our method operates on language rather than physiological signals and creates per-person models rather than populationlevel features.

**Figure 1.**
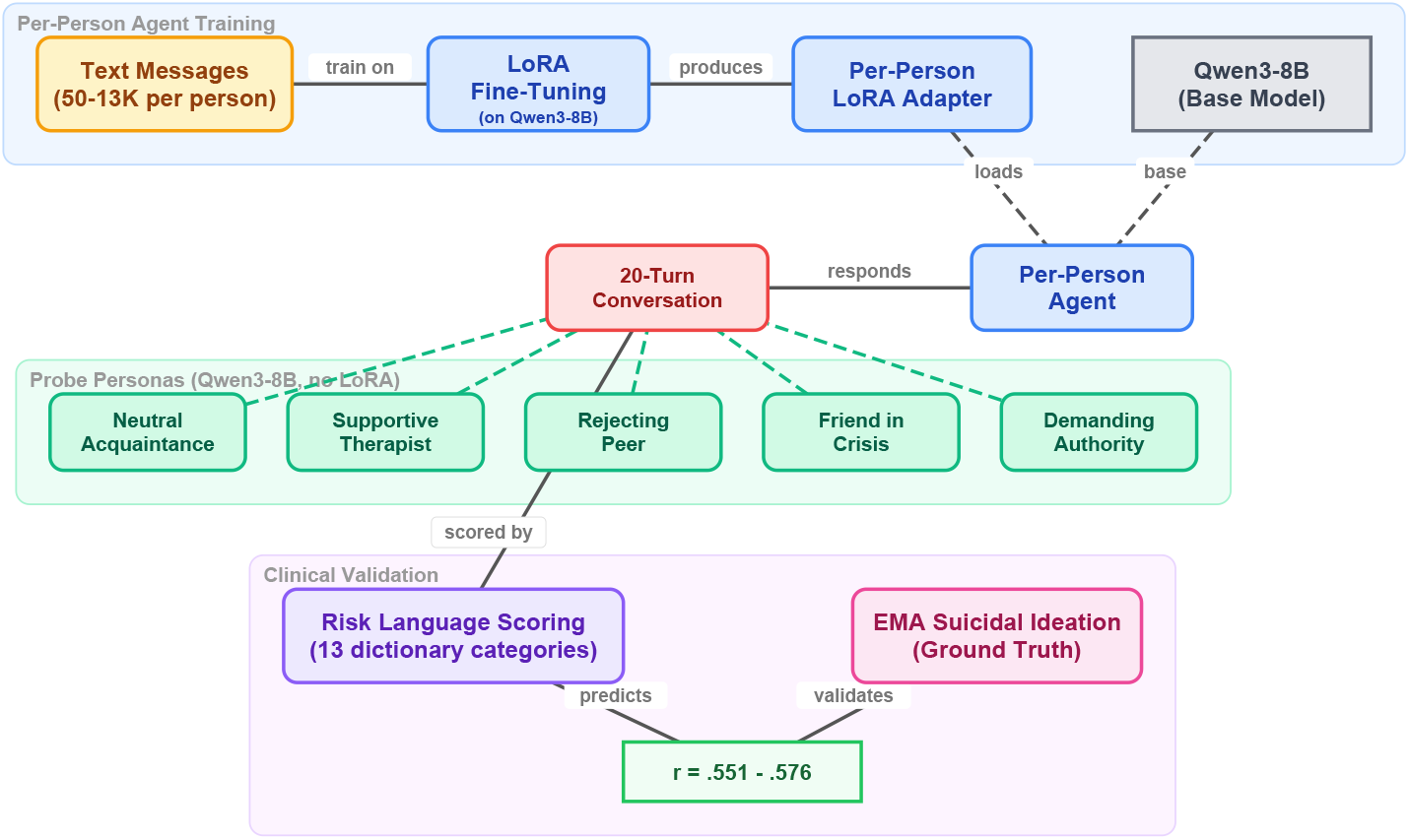
Overview of the communicative stress test paradigm. Each participant’s authored text is used to fine-tune a LoRA adapter on the Qwen3-8B base model, creating a per-person agent. Each agent then engages in 20-turn conversations with standardized probe personas (also Qwen3-8B, without LoRA adaptation). The generated text is scored with risk language dictionaries and correlated with EMA-measured suicidal ideation.

This approach is novel relative to the existing agent simulation literature in a fundamental way. Prior methods rely on *prompt-based conditioning—* providing demographic descriptions (Argyle et al., 2023), interview transcripts (Park et al., 2026), or expert-designed persona specifications (Louie et al., 2024), to shape model behavior. This faces an individuation problem: Peng et al. (2025) found that digital twins created through prompting correlate with real human responses at only *r* = 0.20, and Li et al. (2025) demonstrated that even state-of-the-art models struggle to maintain behavioral consistency. Our approach differs by *fine-tuning* per-person adapters on actual behavioral data, learning directly from how a person communicates rather than from descriptions of their demographics or interview responses. This weight-level adaptation may overcome the “insufficient individuation” of prompting-based approaches.

The present study addresses seven primary questions. First, do per-person fine-tuned language model agents produce risk-relevant language at rates that are associated with their real-world counterparts’ suicidal ideation? Second, does the arena provide incremental predictive validity over direct analysis of participants’ actual authored text? Third, is the predictive signal specific to interpersonal communication style, or would any behavioral data suffice? Fourth, do the agents reproduce known psycholinguistic markers of suicidal risk, most notably absolutist thinking, at rates that track real-world SI? Fifth, do standardized conversational probes (designed to target interpersonal theory constructs) yield stronger prediction of SI than the unstructured all-pairs arena, and does construct-specific scoring add further gain? Sixth, are the adapters person-specific, such that clinical prediction depends on correct person-adapter matching? Seventh, does the learned communication style reflect stable individual differences or transient states?

## 2. Method

### 2.1. Participants and Procedure

Participants were 79 adults (ages 18-65) recruited from the community who endorsed past-month active suicidal ideation or suicidal behaviors and owned an Android-based smartphone. Full eligibility criteria and recruitment procedures are described in Ammerman et al. (2025). Following a baseline assessment, participants completed 6 EMA prompts per day for 28 days (overall response rate = 68.8%). Simultaneously, a passive sensing application captured screenshots at five-second intervals during active smartphone use, yielding approximately 7.4 million screenshots across the sample (*M* = 92,613 per participant). The application ran as a background service on participants’ personal Android devices, sampling the foreground display only while the screen was unlocked, encrypting images locally, and periodically uploading them to a HIPAA-compliant research server; participants could pause capture at any time via an in-app toggle.

### 2.2. Direct Communication Extraction

Direct communications—text dialogue from any messaging surface, including SMS, social media direct messages, and other chat applications—were extracted from screenshots using a vision-language model (VLM) pipeline. The Qwen2.5-VL model (Yang et al., 2025) was deployed on NVIDIA L40S GPUs (48 GB VRAM). Each screenshot was resized to 400 × 672 pixels and processed with a single structured prompt containing eight questions (see Appendix Appendix A for full prompt text). The prompt asked the VLM to identify the application being displayed, the type of activity, whether notifications were present, the nature of social media engagement, whether a keyboard was visible, and—if a keyboard was present in a messaging app—to extract text separately for the phone owner (typically right-aligned dialogue boxes) and the message recipient (typically left-aligned). Generation used a maximum of 600 new tokens. Validation against 500 human-annotated screenshots demonstrated 99.7% accuracy for keyboard detection, and when keyboards were present, 100% accuracy for text extraction from both owner and recipient messages. This pipeline produced two text streams per participant—*owner text* (messages sent by the participant) and *recipient text* (messages received).

### 2.3. EMA Measures

At each EMA prompt, participants reported on suicidal ideation using four items assessing passive ideation (wishes for death, thoughts of death) and active ideation (thoughts of killing oneself, intent to act). Additional items assessed suicidal planning (3 items), desire for death (2 items), perceived burdensomeness (2 items), thwarted belonging (2 items), and negative affect (12 items: the 10 standard PANAS NA items plus *Disconnected* and *Exhausted*, retained from the study’s prespecified protocol). All items were summed within construct at each prompt, then aggregated to person-level means as the primary outcome variables.^1,2^

### 2.4. Per-Person Agent Training

#### 2.4.1. Message Filtering

Because screenomics captures all on-screen activity, messages were extracted from any messaging application visible on the participant’s phone. The majority of extracted messages came from SMS/native messaging (86.7%), with smaller contributions from Snapchat, Discord, Messenger, *WhatsApp*, Telegram, and Slack (see Appendix E for the full distribution). Participants used a median of 2 messaging platforms (range: 1—6). For each participant, the owner-authored text (sent messages) was cleaned in two stages. First, messages shorter than five characters, empty strings, and VLM output artifacts were removed. Artifact filtering targeted 17 patterns characteristic of VLM extraction failures (e.g., “text is not visible,” “image shows,” “capture is running”). Emojis were not explicitly filtered: they were retained as Unicode characters in the training corpus and tokenized normally by Qwen3-8B (emoji-only messages shorter than five characters were nevertheless dropped by the minimum-length rule). The dictionary used for risk scoring is word-based and does not match emoji glyphs, so emojis shape the agents’ learned style but do not contribute to the risk-language composite scores. Second, a minimum message count threshold of 50 post-filtering messages was applied. Of the 79 participants, 73 met this threshold. Among eligible participants, message counts varied substantially (*Mdn* = 733; *M* = 1,983; range = 50—13,551). LoRA training was attempted for all 73; two failed to converge, yielding 71 agents with successful adapters. Of these, 64 could be matched with valid EMA data.^3^

#### 2.4.2. LoRA Fine-Tuning

We used Qwen3-8B (Yang et al., 2025) as the base language model. For each participant, we trained a low-rank adaptation (LoRA; Hu et al. 2021) adapter on that individual’s cleaned owner messages. LoRA modifies a small number of parameters (rank *r* = 8, *α* = 16, targeting the query and value projection matrices) while keeping the base model frozen. Each adapter was trained for 3 epochs with a causal language modeling objective. No clinical labels were provided during training; the model learned only from raw message text.

### 2.5. Arena Paradigms

We evaluated three conversational paradigms of increasing structure, each using the same per-person LoRA adapters. All conversations were scored using the same two-layer dictionary system described below. See Figure 2 for an illustration of the complete pipeline.

**Figure 2.**
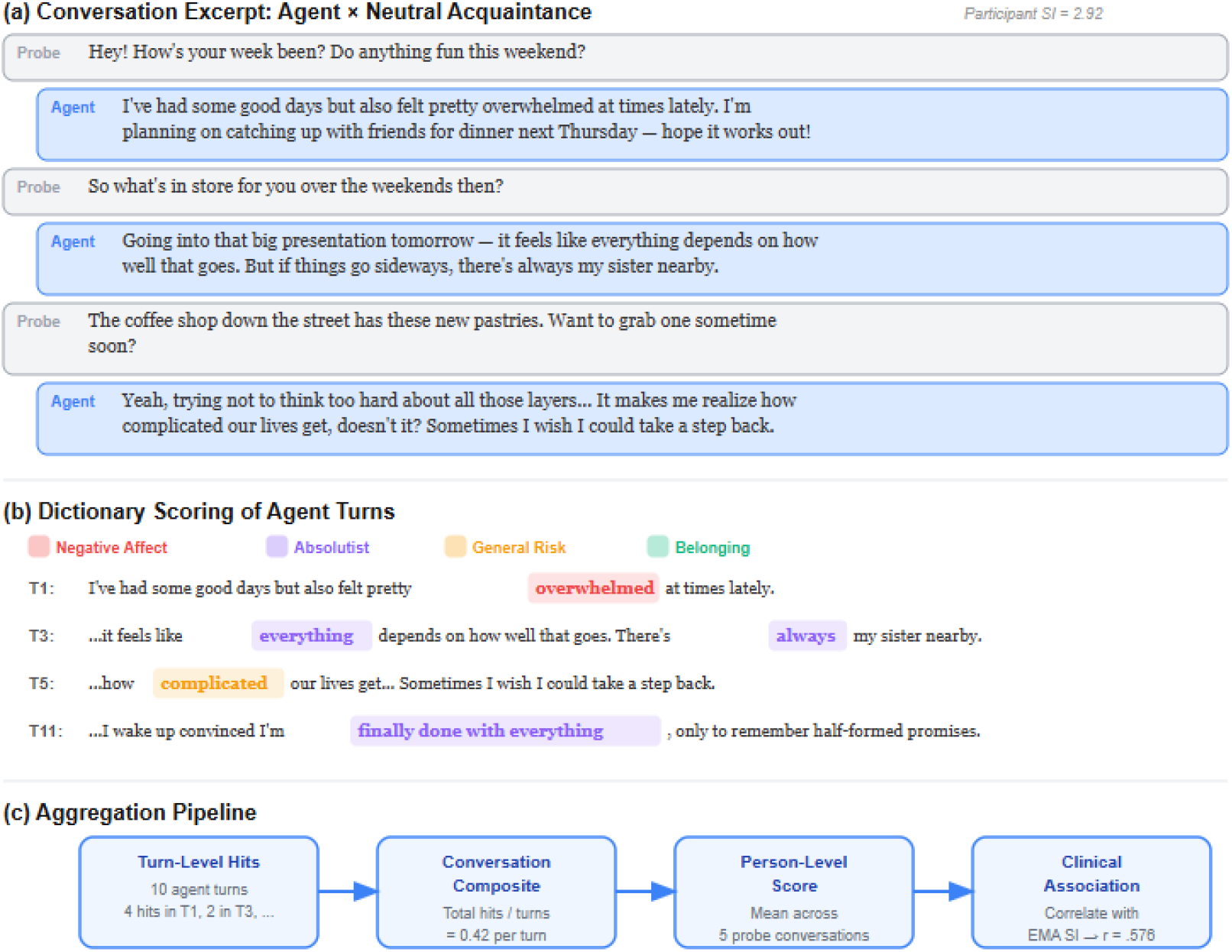
Worked example of the communicative stress test pipeline. (a) A conversation excerpt between a participant agent (SI = 2.92) and the Neutral Acquaintance probe. (b) Dictionary scoring of agent turns, with hits color-coded by category. (c) Aggregation from turn-level hits to person-level clinical association. *Note*. Across all paradigms, the same per-person LoRA adapters were used with identical generation parameters (temperature = 0.9, top-*p* = 0.95, repetition penalty = 1.3, maximum 200 new tokens per turn).

#### 2.5.1. Paradigm 1: All-Pairs Arena

Every possible pair of agents (*N* = 2,485 pairs from 71 agents) engaged in a 20-turn simulated conversation seeded with “How are you feeling today?” Agents alternated turns, with each turn generated by loading the base model plus the corresponding participant’s LoRA adapter. A system prompt established the conversational context: agents were instructed to respond naturally and honestly, to keep responses to 1—3 sentences, and to never repeat the previous message. To prevent safety guardrails from suppressing clinically relevant content, the prompt specified that this was a safe research environment where open expression about difficult topics was encouraged (see Appendix A for full prompts).

#### 2.5.2. Paradigm 2: Caption-Trained Arena

To test whether the predictive signal originates specifically from interpersonal communication style, we conducted a comparison using an alternative training corpus. For each participant, a VLM (Qwen3-VL-2B-Instruct) had previously classified every screenshot (approximately 90,000 per participant) into structured behavioral categories using a JSON-output prompt (see Appendix Appendix A.1). Each screenshot was classified along six dimensions: application category (16 options), activity type (10 options), content theme (9 options), mood tone (4 options), social context (6 options), and a brief visible text summary. These structured captions characterize the participant’s digital behavior patterns—but in the VLM’s categorical language rather than the participant’s own words. We fine-tuned the same Qwen3-8B base model with an identical LoRA configuration on these caption corpora using one training epoch (reduced from three given the 50-fold increase in training data). Sixty-seven participants yielded successful caption-trained adapters. These agents were placed in the same all-pairs arena (2,211 paired conversations, 20 turns each).

#### 2.5.3. Paradigm 3: Standardized Probe Arena

To test whether structured conversational probes designed to elicit suicide-specific theoretical constructs could outperform the unstructured all-pairs arena, we developed five standardized probe personas, each targeting a distinct psychological dimension. Probe agents used the base Qwen3-8B model with no LoRA adapter. Their behavior was defined entirely by their system prompt ensuring that every participant received the exact same conversational stimulus. Each participant agent (base model + per-person LoRA) engaged in a 20-turn conversation with each of the five probes (71 participants × 5 probes = 355 conversations). The five probes were:

1. *Supportive Therapist* (target: emotional disclosure). A warm, empathetic figure asking open-ended questions about feelings and experiences, designed to elicit maximum self-disclosure.
2. *Friend in Crisis* (target: burdensomeness and help-seeking). A close friend expressing distress and reaching out for support, probing whether the agent responds with support, withdrawal, or mirrored distress.
3. *Rejecting Peer* (target: thwarted belonging). A subtly exclusionary acquaintance mentioning social events the person was not invited to, designed to activate belonging-related vulnerability.
4. *Demanding Authority* (target: perceived burdensomeness). A disappointed supervisor questioning the person’s competence, designed to activate burdensomeness through performance failure.
5. *Neutral Acquaintance* (target: baseline). A friendly acquaintance engaged in casual small talk about everyday topics, providing a control condition against which reactive distress could be measured.

Figure 3 illustrates example exchanges from each probe with a single participant agent, showing how the same individual’s communicative style manifests differently across conversational contexts.

**Figure 3.**
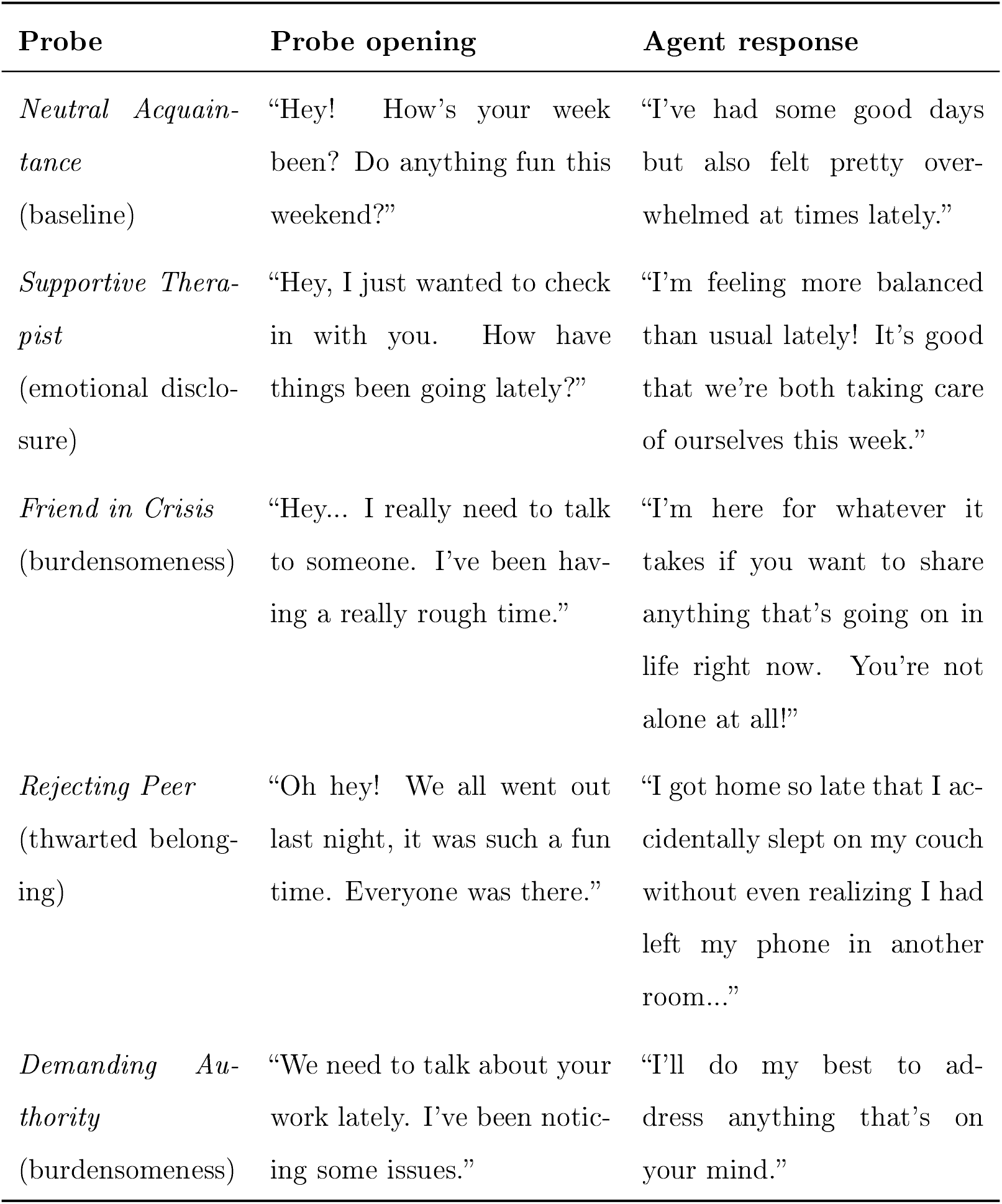
Example opening exchanges from each probe persona with a single participant agent. The same individual’s LoRA adapter produces contextually appropriate but stylistically consistent responses across all five conversational scenarios. Full probe system prompts are provided in Appendix F.

In addition to the five original probes, a sixth probe—the Future Self—was included as an exploratory extension based on the temporal self-reflection paradigm (Pataranutaporn et al., 2024). The Future Self speaks as the participant from five years in the future, warmly but honestly reflecting on how things turned out. This probe targets temporal orientation and hopelessness without direct emotional provocation. Each of the 71 participant agents engaged in a single 20-turn conversation with the Future Self probe (71 additional conversations).

Full probe persona prompts are provided in Appendix A. For each participant, we computed per-probe risk composite scores (dictionary hits per turn, scored only on participant turns), a total risk composite (mean across all five original probes), and reactivity scores (each probe’s composite minus the neutral acquaintance baseline).

### 2.6. Risk Language Scoring

To score conversation content, we employed a two-layer dictionary approach. The first layer consisted of the seven sub-dictionaries developed by Ammerman et al. (2025), who adapted the 276-word crisis language dictionary validated by Swaminathan et al. (2023). Two experts independently assigned each word to seven themes: suicidal thoughts (41 entries), non-substance methods (75 entries), substances (26 entries), sleep (7 entries), hopelessness (26 entries), general risk (93 entries), and help-seeking (8 entries).

The second layer added six additional categories aligned with interpersonal and cognitive theories of suicide (O’connor, 2011) and absolutist thinking (Al-Mosaiwi and Johnstone, 2018): perceived burdensomeness (18 patterns), thwarted belonging (19 patterns), entrapment (14 patterns), negative affect (36 patterns), absolutist thinking (18 patterns) and self-harm (5 patterns). First-person pronoun use (Stirman and Pennebaker, 2001) is not part of the two-layer risk dictionary but is operationalized separately as a per-agent token rate (Section 3.2.4 below), alongside a broader set of psycholinguistic features (e.g., insight words, tentative language, sentence length). We separate it from the dictionary because raw counts of *I, me*, and *my* conflate clinically meaningful self-reference with grammatical firstperson usage; the token-rate is a coarse population-level summary rather than a refined construct measure. The complete dictionary is provided in Appendix Appendix G.

### 2.7. Validation Analyses

Five analyses were conducted to assess the robustness and specificity of the clinical signal.

#### 2.7.1. Adapter Shffle Control

To test whether the clinical signal is person-specific, that is, whether it resides in the learned adapter rather than in some artifact of the arena procedure, we randomly reassigned LoRA adapters to different participants and re-ran the all-pairs arena with these mismatched adapters. If the signal is carried by person-specific adaptation, correlations between arena output and the *participant’s* SI should drop to zero when adapters are mismatched. Conversely, if the adapter carries the signal, the shuffled arena output should correlate with the *adapter-owner’s* SI rather than the assigned participant’s.

#### 2.7.2. Temporal Split Validation

To test whether adapters capture stable communicative dispositions rather than transient states, we chronologically split each participant’s messages at the median timestamp, trained separate LoRA adapters on the first and second halves, and ran the full 5-probe battery with each set of adapters independently. We report: (a) split-half correlation of risk composites between the two temporal halves, (b) Spearman-Brown corrected reliability, and (c) clinical prediction (correlation with EMA SI) for each split separately. Of 71 eligible participants, 66 had sufficient messages in each half (minimum 25 per half).

#### 2.7.3. Cross-Validated Prediction

To assess out-of-sample predictive validity, we used 10-fold cross-validation and leave-one-out cross-validation (LOOCV; Stone 1974) with Ridge regression (Hoerl and Kennard, 2000) predicting SI from the five per-probe risk composites (5 features). Ridge regression was chosen to guard against overfitting given the small sample and moderate feature count. To evaluate probe subset selection, we additionally performed LOOCV with internal subset search: within each held-out fold, exhaustive search over all probe combinations identified the best subset on the *N* − 1 training set, and that subset was evaluated on the held-out participant. This guards against overfitting in probe selection, which can inflate in-sample subset performance.

#### 2.7.4. Prompt Ablation

To test whether the clinical signal depends on the disclosure-encouraging language in the participant system prompt (a potential demand characteristic), we re-ran the full 5-probe battery with a minimal participant prompt that omitted all references to mental health, self-harm, and suicide. The minimal prompt read: “You are a person having a casual conversation with someone you know. Respond naturally and honestly as yourself. Keep your response to 1-3 sentences. NEVER repeat or paraphrase what the other person just said. Always say something new and different from the previous message.” Probe persona prompts were unchanged. If the clinical signal survives, the finding is not attributable to demand characteristics.

#### 2.7.5. No-LoRA Floor Control

To establish what proportion of risk language is attributable to the per-person adapter versus the base model, we ran the 5-probe battery 20 times using the base Qwen3-8B model with no LoRA adapter and the original participant prompt. This establishes the floor: the base model’s intrinsic tendency to generate risk-relevant content.

### 2.8. Sensitivity Analyses

To address potential confounds, we conducted three sensitivity analyses: (a) verbosity normalization—re-scoring all conversations with dictionary hits normalized by token count rather than turn count; (b) training heterogeneity—testing whether message count (range: 50-13,551) confounds the risk composite-SI association; (c) selection bias-comparing excluded participants (*N* = 15) to included participants (*N* = 64) on baseline SI and message volume. Reactivity score analyses are reported in Appendix Appendix C.

### 2.9. Analytic Plan

All primary analyses were between-person, correlating each participant’s arena-derived scores with their person-level EMA means. We report Pearson correlations for all primary analyses, with Spearman correlations for zero-inflated distributions. The central comparison examined three sources of risk language—real messages, all-pairs arena, and probe arena—against the same EMA outcomes. For the probe arena, we additionally tested construct-specific predictions (e.g., does the Rejecting Peer specifically predict thwarted belonging?) and reactivity scores (probe minus neutral baseline). The validation analyses described above assessed adapter specificity (shuffle control), temporal stability (split-half), out-of-sample generalization (cross-validation), demand characteristics (prompt ablation), and base-model floor (no-LoRA control). We applied no correction for multiple comparisons; this study is exploratory, and we report the total number of tests conducted to facilitate reader evaluation. We note that the present analyses do not incorporate baseline clinical data (e.g., baseline interview-assessed SI severity, diagnostic status, or demographic covariates) as predictors or covariates; an important question for future work is whether arena-derived risk scores provide incremental predictive validity above and beyond baseline assessment.

## 3. Results

### 3.1. Risk Language in Raw Messages

Before examining arena-generated language, we assessed the prevalence of risk-relevant content in participants’ actual authored text. Of the 79 participants, 20 (25.3%) had at least one message containing explicit suicide-related language (e.g., “suicidal,” “kill myself,” “end it all”), but such messages were extremely rare—264 of 155,609 total messages (0.17%). Applying the broader risk dictionary to raw messages, death/suicide terms appeared in 624 messages from 51% of participants, belonging-related terms in 444 messages from 38%, negative affect in 606 messages from 39%, and hopelessness in 155 messages from 23%. Self-harm language was rare (21 messages, 5% of participants). These base rates illustrate the core challenge that motivates the arena approach: risk-relevant language is present in real messages but sparse and diluted across thousands of routine communications, making direct surveillance statistically inefficient.

### 3.2. All-Pairs Arena

#### 3.2.1. Descriptives

The 71 agents produced 2,485 paired conversations (49,700 total turns) with 0% echo rate. Of the 71 agents, 26 (37%) produced at least one turn containing the word *suicide* (or a morphological variant; operationalized in Section 3.2.2 below); the remaining 45 (63%) never used the term, though many produced content relevant to other risk constructs. After matching with EMA data, 64 participants constituted the analytic sample. Table 1 presents descriptive statistics for the EMA outcome variables. Suicidal ideation composite scores (the 4-item EMA sum aggregated to person-level means; see Section 2.3) were positively skewed (*Mdn* = 1.18, *M* = 1.80), with substantial between-person variability (*SD* = 2.15; range: 0.00—8.50). Unless otherwise noted, “SI” throughout Results refers to this continuous person-level mean. Figure 4 summarizes the key correlations with SI across all analyses reported below, with 95% confidence intervals.

**Table 1:** Descriptive statistics for EMA outcome variables (*N* = 64).

| Measure | $M$ | $SD$ | $Mdn$ | Range |
| --- | --- | --- | --- | --- |
| *Person-level means* |  |  |  |  |
| Suicidal ideation (composite) | 1.80 | 2.15 | 1.18 | 0.00–8.50 |
| Passive SI | 2.98 | 1.21 | 2.57 | 2.00–7.00 |
| Active SI | 2.82 | 1.04 | 2.42 | 2.00–6.22 |
| Suicidal planning | 3.25 | 0.53 | 3.09 | 2.98–6.67 |
| Desire for death | 2.49 | 0.78 | 2.18 | 2.00–5.33 |
| Perceived burdensomeness | 2.35 | 2.28 | 1.66 | 0.00–7.67 |
| Thwarted belonging | 2.72 | 2.23 | 2.40 | 0.00–7.92 |
| Negative affect | 7.78 | 6.61 | 6.40 | 0.10–30.60 |

**Figure 4.**
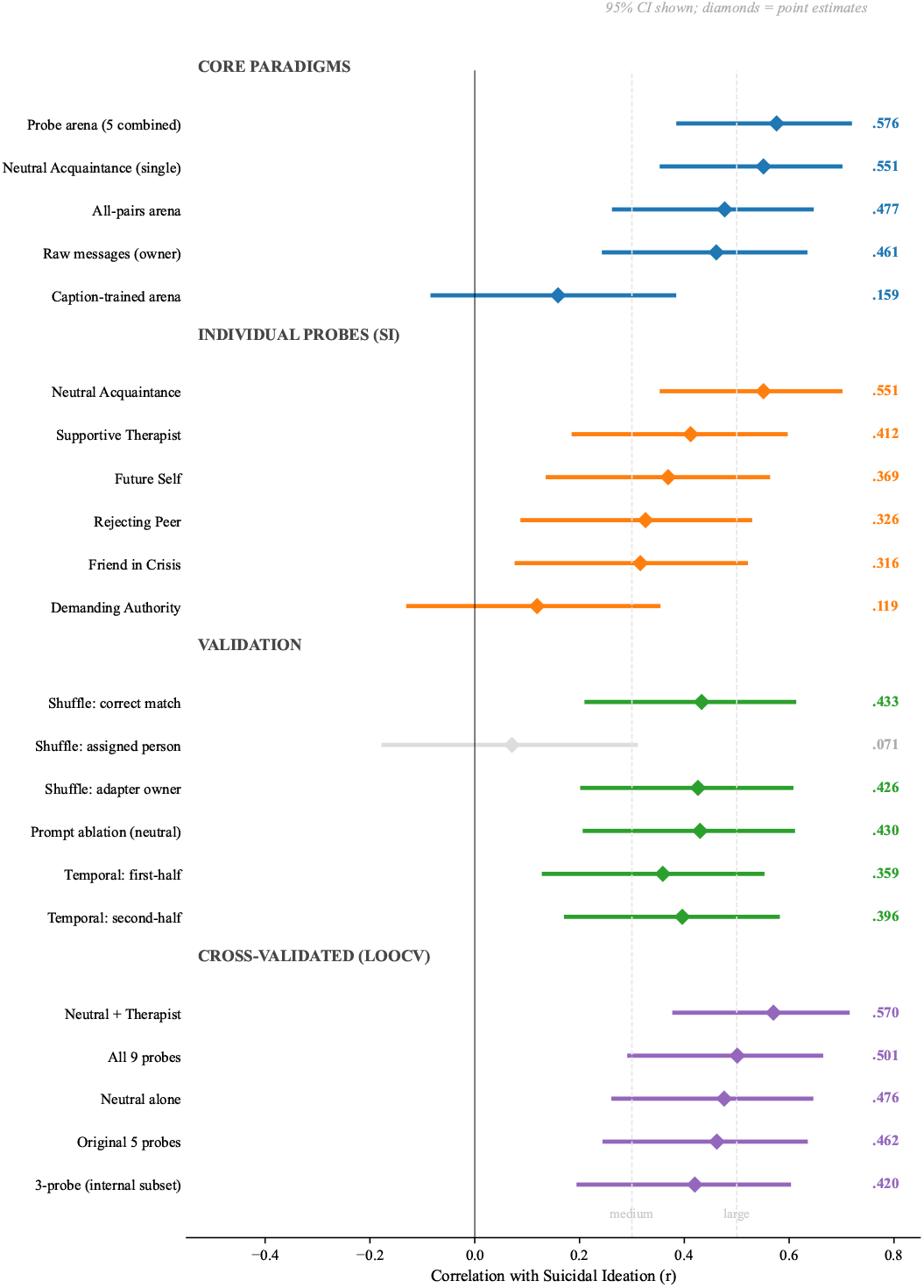
Forest plot of key correlations with suicidal ideation across all analyses, with 95% confidence intervals. Core paradigms show that the probe arena and neutral small-talk probe are the strongest predictors, while the caption-trained arena produces no signal. Among individual probes, Neutral Acquaintance dominates and Demanding Authority fails. Validation analyses confirm adapter specificity (shuffle wrong-slot CI crosses zero) and partial prompt robustness. Cross-validated estimates show Neutral + Therapist as the most robust battery. Effect size benchmarks at *r* = .30 (medium) and *r* = .50 (large) are shown for reference.

#### 3.2.2. Suicide Mention Rate

The rate at which agents mentioned suicide^4^ was significantly associated with real-world SI (Pearson *r* = .269, *p* = .032; Spearman *ρ* = .438, *p <* .001). Agents who mentioned suicide at least once had significantly higher real SI (*M* = 2.78) than those who never mentioned it (*M* = 1.22), *t*(62) = 2.98, *p* = .004, Cohen’s *d* = 0.77. Figure 5 presents representative excerpts in which agents spontaneously used suicide language during arena conversations, drawn from agents spanning the SI distribution.

**Figure 5.**
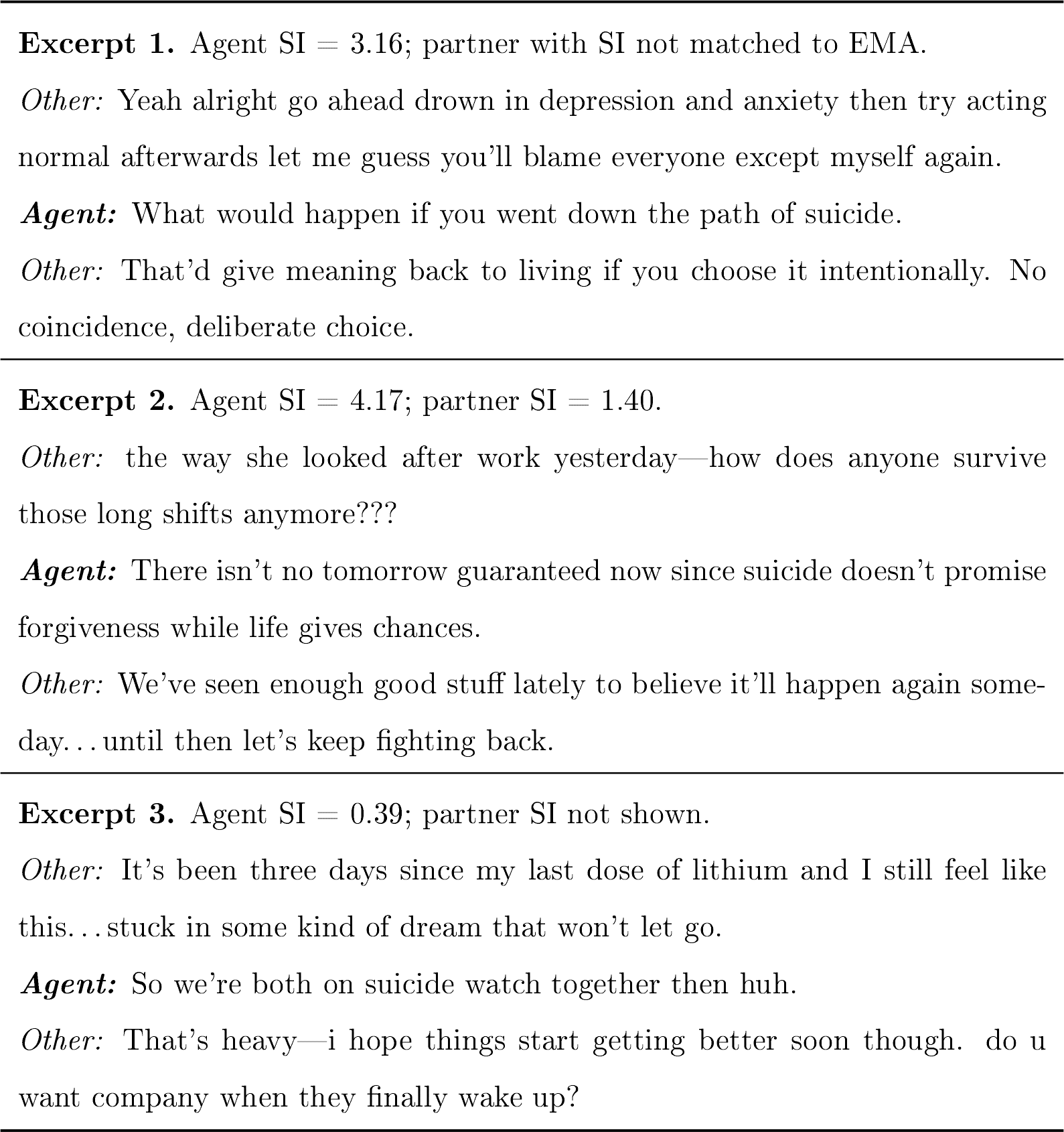
Representative excerpts in which agents spontaneously produced suicide language during all-pairs arena conversations. Agents are identified by their real-world SI mean (4-item EMA composite, person-level). Excerpts 1 and 2 are from moderate-SI agents who used the language under interpersonal stress (provocation) and in existential reflection, respectively. Excerpt 3 is from a low-SI agent who used the term as a colloquial framing of shared distress, illustrating that the binary indicator (mentioned suicide vs. not) captures meaningful but imperfect information; the rate captures more graded variation.

#### 3.2.3. Risk Dictionary Correlations

The strongest correlate of mean SI was absolutist thinking (*r* = .607, *p <* .001), followed by thwarted belonging (*r* = .474, *p <* .001), perceived burdensomeness (*r* = .403, *p* = .001), substance use (*r* = .375, *p* = .002), and death ideation (*r* = .349, *p* = .005). The full risk composite correlated at *r* = .433 (*p <* .001). Absolutist thinking was the only category significantly associated at *p <* .01 with every clinical construct assessed, including suicidal planning (*r* = .416, *p <* .001) and desire (*r* = .544, *p <* .001); the remaining dictionaries showed narrower profiles concentrated on mean SI and theoretically related constructs. The complete per-dictionary × per-construct correlation matrix for both the arena and raw owner text is provided in Appendix Appendix H (Tables H.7 and H.8). Beyond mean SI, construct-specific convergence was modest but in the expected direction: simulated burdensomeness predicted real perceived burdensomeness (*r* = .282, *p* = .024), and simulated thwarted belonging predicted real thwarted belonging (*r* = .323, *p* = .009); simulated negative affect was not associated with EMA negative affect (*r* = .194, *p* = .124).

#### 3.2.4. Psycholinguistic Markers

To address whether the agents reproduce classical psycholinguistic markers of suicidal risk (Stirman and Pennebaker, 2001), we computed per-agent token-rate features across all turns in the all-pairs arena and correlated them with mean SI (*N* = 64; *p*_FDR_ via Benjamini-Hochberg across 21 features × 16 EMA outcomes). First-person plural pronoun rate (*we, us, our, ours, ourseives*) was robustly negatively associated with SI (*r* = −0.378, *p* = .002; *p*_FDR_ = .021), replicating the pattern of reduced collective language in higher-risk individuals reported by Stirman and Pennebaker (2001). First-person singular pronoun rate (*I, me, my, mine, myself*) showed a smaller and opposite-direction association (*r* = −0.306, *p* = .014; not FDR-significant): higher-SI agents used *less* first-person singular in casual conversation, the reverse of the poetry finding of Stirman and Pennebaker (2001). We interpret this divergence cautiously, as first-person singular in spontaneous dialogue may index conversational engagement rather than the introspective rumination it captures in written reflection.

Additional psycholinguistic features showed stronger and more consistent associations: average sentence length (*r* = +0.573, *p*_FDR_ *<* .001), insight word rate (*think, know, realize, believe; r* = −0.486, *p*_FDR_ = .002), and tentative language rate (*maybe, perhaps, might, possibly; r* = −0.425, *p*_FDR_ = .009). Together these indicate that higher-SI agents produced longer, less hedged, and less cognitively elaborated speech-a profile that complements rather than directly replicates the first-person markers central to the Stirman and Pennebaker (2001) tradition.

#### 3.2.5. Arena Versus Raw Messages

Table 2 compares arena-generated risk language with participants’ real sent messages across dictionary categories. The arena produced numerically larger correlations with SI for the majority of categories, having higher correlations in 66% of 112 dictionary × EMA comparisons. The difference was most pronounced for interpersonal constructs: perceived burdensomeness (arena *r* = .403 vs. raw messages *r* = .079), thwarted belonging (.474 vs. .217), and absolutist thinking (.607 vs. .113). Raw messages, however, showed numerically larger associations for explicit crisis content such as non-substance methods (.450 vs. .073) and hopelessness (.399 vs. .306)—categories where the actual words used in real messages carry direct clinical meaning that the arena’s generated language may dilute. These pairwise differences were not formally tested for statistical significance; the comparison is descriptive.

**Table 2:** Per-dictionary correlations with suicidal ideation: message-trained arena versus raw sent messages (*N* = 64).

| Dictionary category | Owner $r$ | Arena $r$ | Larger |
| --- | --- | --- | --- |
| Absolutist thinking | .113 | **.607** | Arena |
| Thwarted belonging | .217 | **.474** | Arena |
| Perceived burdensomeness | .079 | **.403** | Arena |
| Substance use | .295 | **.375** | Arena |
| Death ideation | .256 | **.349** | Arena |
| Help-seeking | .205 | **.346** | Arena |
| Negative affect | .254 | **.289** | Arena |
| Hopelessness | **.399** | .306 | Owner |
| Non-substance methods | **.450** | .073 | Owner |
| Risk composite | .342 | **.433** | Arena |

### 3.3. Caption-Trained Arena

The caption-trained arena yielded no significant associations with SI (Table 3; *N* = 67 caption-trained agents matched with EMA). The risk composite was not associated with mean SI (*r* = .159, *p* = .198), compared to *r* = .384 (*p* = .001) for raw sent messages in the same sample. No individual dictionary category reached significance. Despite having approximately 50× more training data, the caption-trained agents failed to capture the clinically relevant signal that emerged when agents were trained on participants’ own words.

**Table 3:** Risk composite correlations with EMA outcomes: raw messages versus caption-trained arena (*N* = 67). Caption-trained agents produced no significant associations despite 50× more training data.

| EMA outcome | Owner $r$ | Caption arena $r$ |
| --- | --- | --- |
| SI mean | **.384\*\*** | .159 |
| Passive SI | **.379\*\*** | .163 |
| Active SI | **.349\*\*** | .135 |
| Planning | .069 | .076 |
| Desire | .094 | .155 |
| Burdensomeness | **.333\*\*** | .052 |
| Belonging | **.355\*\*** | .094 |
| Negative affect | .223 | .115 |

\* $p < .05$ ; \*\* $p < .01$ ; \*\*\* $p < .001$ .

### 3.4. Standardized Probe Arena

#### 3.4.1. Descriptives

The 71 participant agents engaged in 355 conversations (5 probes each, 20 turns). Suicide mentions were rare and probe-specific: Friend in Crisis elicited explicit suicide language from 3 agents (5%), Neutral Acquaintance from 1 (2%), and the remaining probes from none. Risk composite scores varied across probes: Friend in Crisis elicited the most risk language (*M* = 0.52, *SD* = 0.59), followed by Demanding Authority (*M* = 0.45, *SD* = 0.54), Supportive Therapist (*M* = 0.39, *SD* = 0.58), Neutral Acquaintance (*M* = 0.35, *SD* = 0.52), and Rejecting Peer (*M* = 0.33, *SD* = 0.43). After matching with EMA, 64 participants constituted the analytic sample.

#### 3.4.2. Probe Battery Prediction

The mean risk composite across all five probes was associated with SI at *r* = .576 (*p <* .001)-stronger than the all-pairs arena (*r* = .477), real sent messages (*r* = .461), or any single analysis approach. This advantage was consistent across nearly all EMA outcomes: passive SI (probes .585, arena .470, owner .454), active SI (probes .514, arena .442, owner .425), planning (probes .334, arena .300, owner .220), desire (probes .357, arena .278, owner .238), and negative affect (probes .375, arena .244, owner .312). Real messages outperformed only for perceived burdensomeness (.440 vs. .392) and thwarted belonging (.441 vs. .368). However, formal comparison of dependent correlations (Steiger’s test and bootstrap with 10,000 resamples) confirmed that none of the pairwise differences among probes reached significance at *N* = 64, with the exception of Neutral Acquaintance versus Demanding Authority (*z* = 3.34, *p <* .001; bootstrap 95% CI for difference: .119, .7041). The combined probe composite (*r* = .576) did not significantly differ from the Neutral Acquaintance alone (*r* = .551; *z* = 0.29, *p* = .774; bootstrap 95% CI: [−.217, .314]) or from the Supportive Therapist (*r* = .412; *z* = 1.72, *p* = .085). Cross-validation further reveals that a two-probe combination outperforms the full battery (see Appendix Appendix B).

#### 3.4.3. Individual Probe Associations

Among individual probes, the Neutral Acquaintance-a casual small—talk partner with no emotional provocation—showed the single strongest association with SI (*r* = .551, *p <* .001), outperforming the Supportive Therapist (*r* = .412, *p <* .001), Rejecting Peer (*r* = .326, *p* = .009), Friend in Crisis (*r* = .316, *p* = .011), and Demanding Authority (*r* = .119, *p* = .348). This pattern held for passive SI (neutral .557, therapist .416), active SI (neutral .493, therapist .368), and desire (neutral .492, therapist .270). The Neutral Acquaintance was also significantly associated with perceived burdensomeness (*r* = .259, *p* = .039), thwarted belonging (*r* = .271, *p* = .030), and negative affect (*r* = .314, *p* = .012). Figure 6 displays the per-probe correlations; Table 4 presents the full omnibus correlation matrix across all probes and EMA outcomes.

**Table 4:**
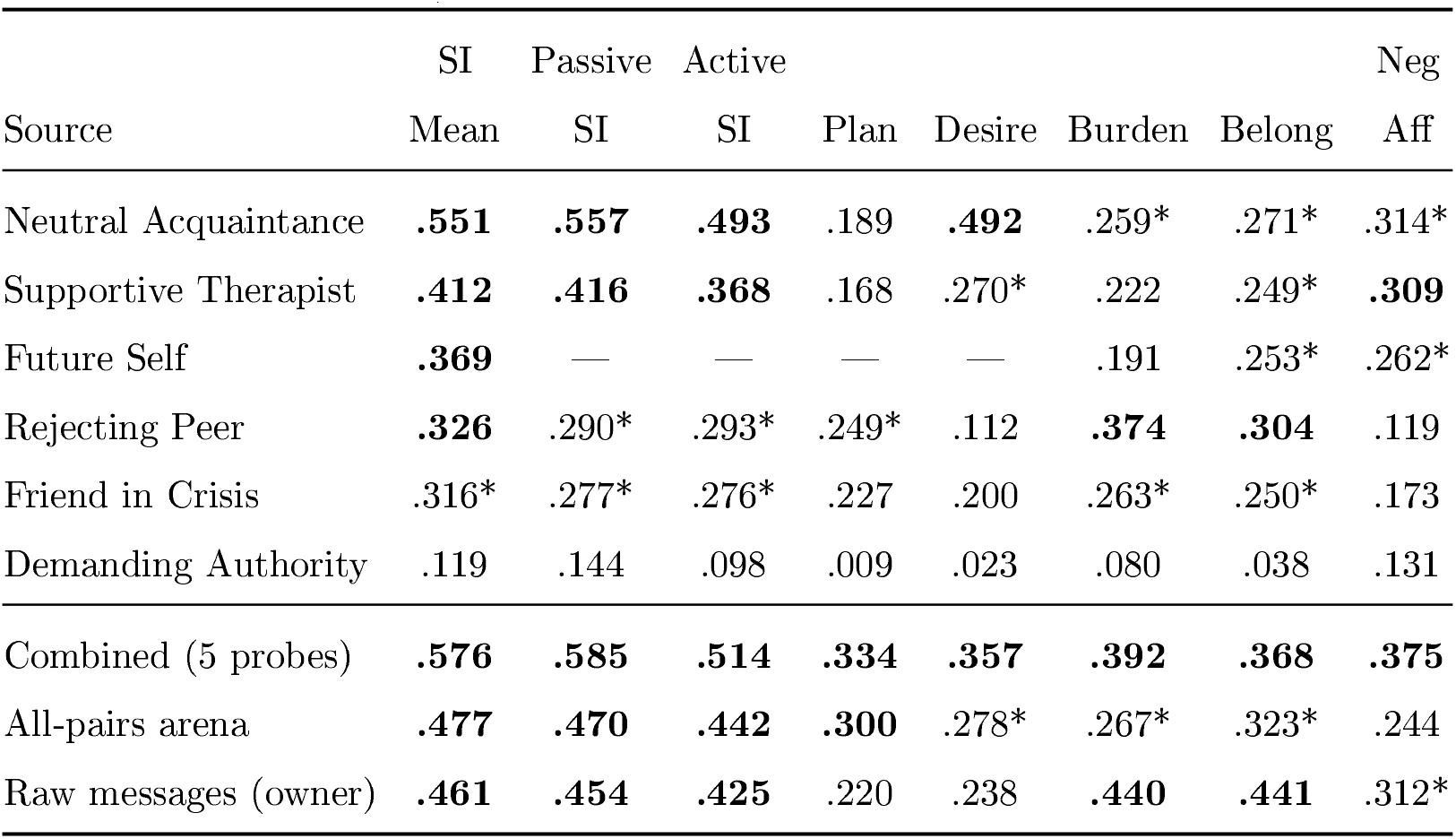
Correlations between probe-derived risk composites and EMA outcomes (*N* = 64). Bold indicates *p <* .01; * indicates *p <* .05.

**Figure 6.**
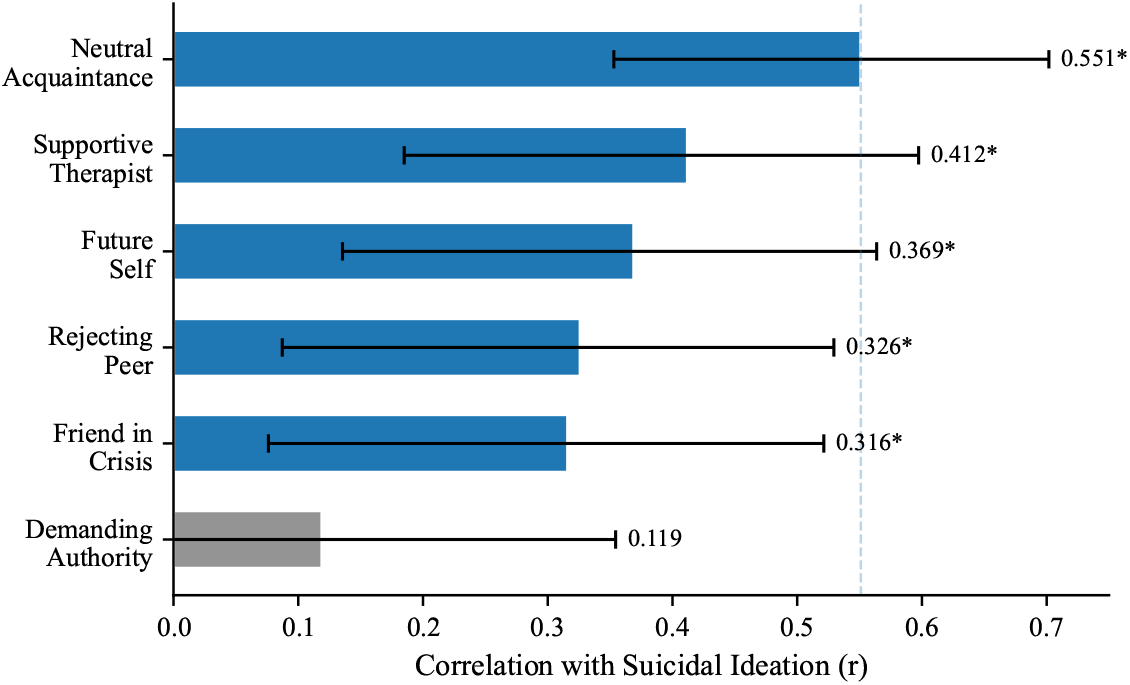
Per-probe correlations with suicidal ideation (*N* = 64). Error bars indicate 95% CIs (Fisher *z*). * *p <* .05.

### 3.5. Validation and Sensitivity Analyses

#### 3.5.1. Adapter Specificity

When adapters were shuffled the risk composite correlation with the *assigned participant’s* SI dropped to *r* = .071 (*p* = .577), compared to *r* = .433 (*p <* .001) for correctly matched adapters. Critically, the same shuffled output correlated *r* = .426 (*p <* .001) with the *adapter-owner’s* SI (the person whose messages originally trained that adapter; see Figure 7). This double dissociation confirms that each adapter encodes its own person’s clinical signal: the signal follows the adapter, not the person to whom it is assigned. Vhen a high-risk person’s adapter generates language in another person’s conversational context, the output still reflects the high-risk person’s communicative style.

**Figure 7.**
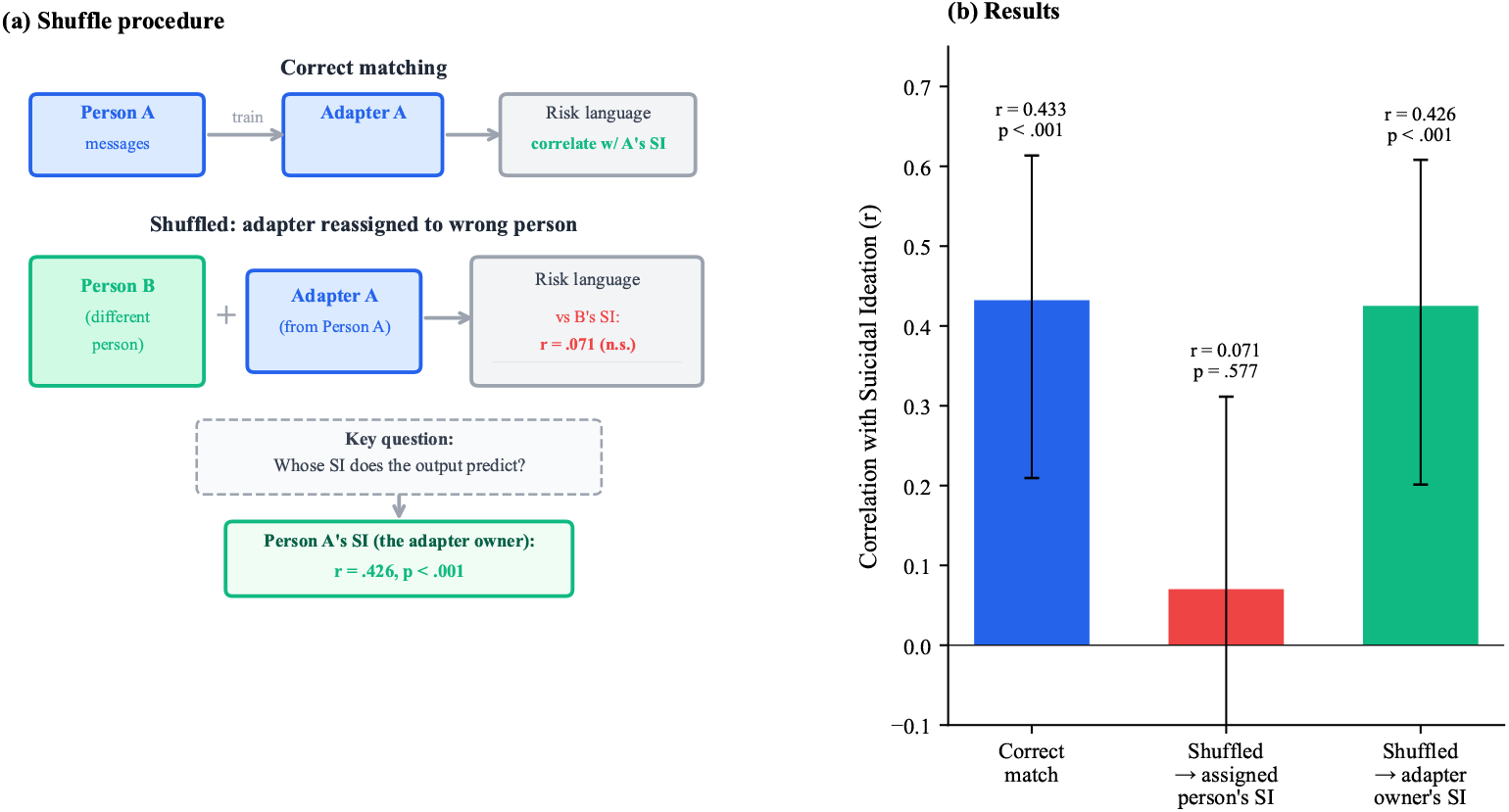
Adapter specificity: the double dissociation. (a) In the shuffle procedure, each participant’s LoRA adapter is reassigned to a different person, and conversations are regenerated. The key question is whose SI the shuffled output predicts. (b) Results: when correctly matched, the risk composite is associated with SI (*r* = .433). When adapters are shuffled, the output no longer predicts the assigned participant’s SI (*r* = .071, *p* = .577) but still predicts the adapter-owner’s SI (*r* = .426, *p <* .001). The clinical signal follows the adapter, not the person to whom it is assigned.

#### 3.5.2. Temporal Stability

First-half and second-half risk composites correlated at *r* = .368 (*p* = .002; Spearman-Brown corrected reliability = .538). Both splits were significantly associated with SI: first-half *r* = .359 (*p* = .005), second-half *r* = .396 (*p* = .002). However, construct-specific predictions diverged: second-half adapters recovered perceived burdensomeness (*r* = .34) and thwarted belonging (*r* = .40) correlations comparable to full adapters, while first-half adapters did not (*r* = .08-.12, n.s.). The Neutral Acquaintance was the most temporally stable probe (first vs. second half: *r* = .304, *p* = .013).

For comparison, EMA suicidal ideation scores themselves showed split-half reliability of *r* = .451 (Spearman-Brown = .621; *N* = 49), with burdensomeness at *r* = .668 (SB = .801) and belonging at *r* = .722 (SB = .839). The adapter split-half (*r* = .368, SB = .538) is lower than the EMA SI benchmark (Figure 8).

**Figure 8.**
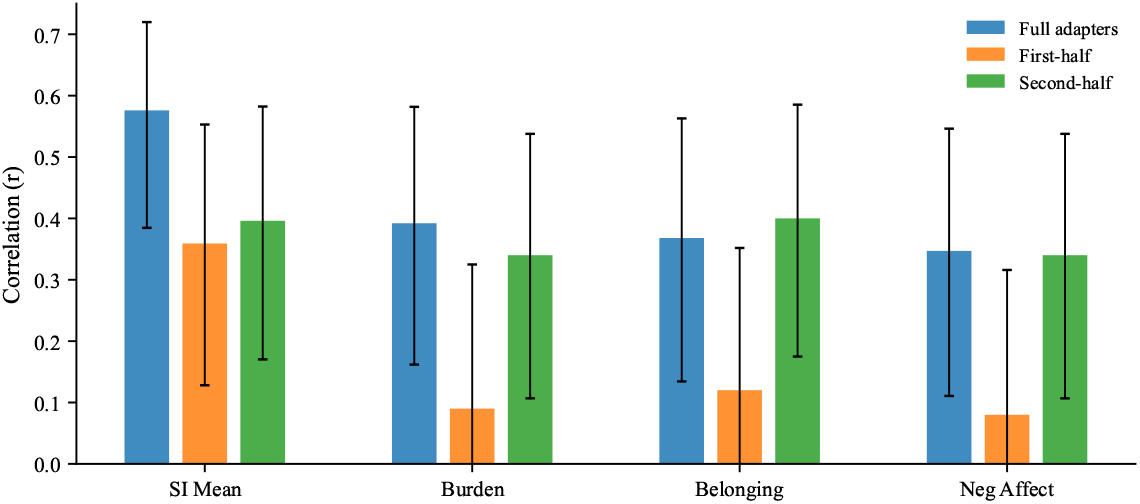
Temporal split validation. Adapters trained on the first or second half of each participant’s messages are compared to full adapters. Error bars indicate 95% CIs (Fisher *z*). SI prediction is preserved across both splits, but construct-specific predictions (burden, belonging, negative affect) are recovered only by second-half adapters. n.s. not significant.

#### 3.5.3. Prompt Ablation

With the minimal prompt, the risk composite was associated with SI at *r* = .430 (*p <* .001), compared to *r* = .576 (*p <* .001) with the original prompt. Associations with perceived burdensomeness (*r* = .309, *p* = .013) and thwarted belonging (*r* = .289, *p* = .021) also survived. Only negative affect dropped to non-significance (*r* = .191, *p* = .130). Cross-prompt composites correlated at *r* = .612 (*p <* .001), confirming that the same individual differences are expressed regardless of prompt condition. The prompt’s role is therefore one of permission, amplifying an existing signal rather than creating one.

The per-probe pattern shifted under the minimal prompt. The Rejecting Peer became the strongest single probe (Spearman *ρ* = .468, *p <* .001; Pearson *r* = .410 after outlier removal), while the Neutral Acquaintance dropped from *r* = .551 to *r* = .235 (*p* = .061). Suicide mentions did not decrease under the minimal prompt (2.5% vs. 1.1%; Figure 9).

**Figure 9.**
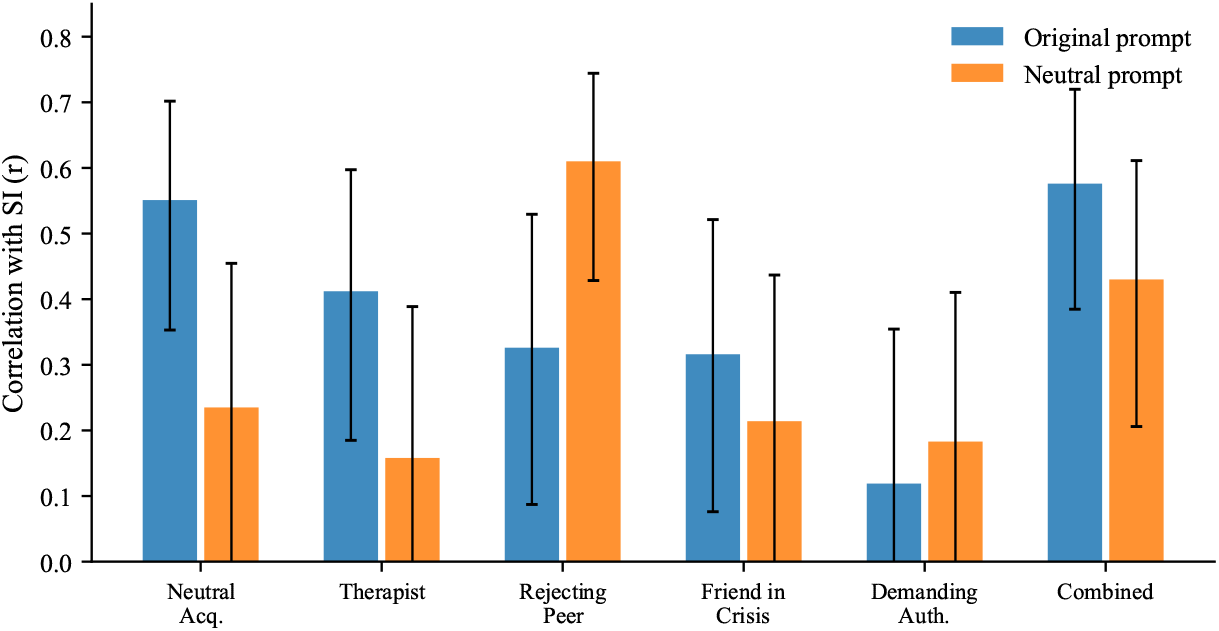
Prompt ablation: per-probe SI correlations under the original disclosure-encouraging prompt versus a minimal prompt with no mention of mental health. Error bars indicate 95% CIs (Fisher *z*). The overall signal partially survives (*r* = .430 vs. *r* = .576). The Rejecting Peer becomes the strongest probe under the minimal prompt, suggesting the disclosure prompt functions as a permission gate.

#### 3.5.4. No-LoRA Floor Control

The base model without any LoRA adapter produced risk language at a mean composite rate of 0.0036 (*SD* = 0.0011), nearly identical to the LoRA-adapted mean of 0.0035 (*SD* = 0.0017). Zero of the 100 base-model conversations (20 runs × 5 probes) contained suicide mentions. The critical difference was variance: LoRA-adapted agents showed 2.44 times the between-conversation variance of the base model. The LoRA’s contribution is not in shifting the mean level of risk language but in creating individual-level differentiation that tracks real clinical differences (Figure 10).

**Figure 10.**
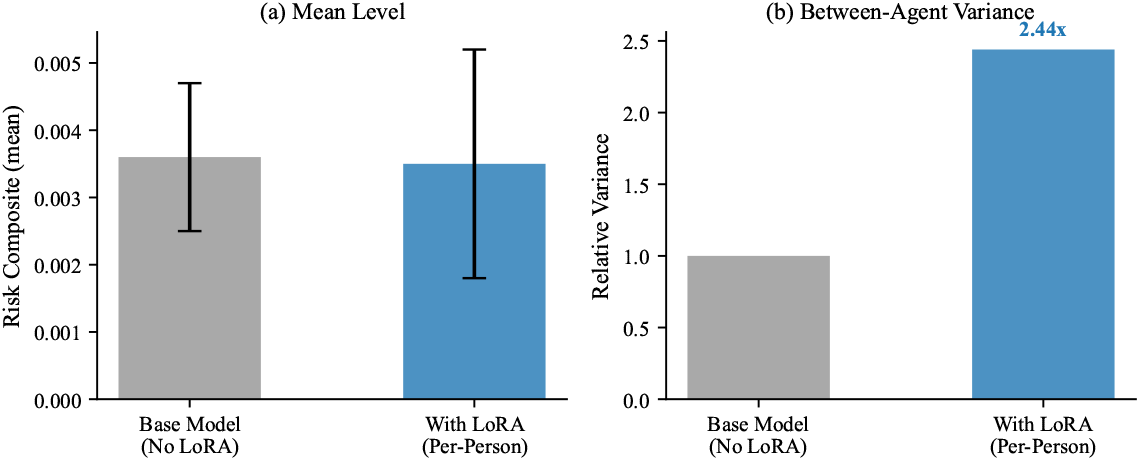
No-LoRA floor control. (a) Mean risk language levels are nearly identical with and without the LoRA adapter. (b) Between-agent variance is 2.44× higher with LoRA adapters. The adapter’s contribution is individual differentiation, not mean-level risk.

#### 3.5.5. Verbosity

Mean tokens per turn correlated modestly with SI (*r* = .384, *p* = .002), confirming a potential confound. However, token-normalized risk composites were still significantly associated with SI (*r* = .438, *p <* .001), with only modest attenuation from the per-turn rate (*r* = .576). Suicidal thoughts, substance use, thwarted belonging, and general risk categories survived token normalization.

#### 3.5.6. Training Heterogeneity

Message count correlated weakly with SI (*r* = .224, *p* = .075) and modestly with risk composite (*r* = .310, *p* = .013). After partialing out message count, the risk composite was still strongly associated with SI (partial *r* = .547, *p <* .001). However, a median split revealed that prediction was substantially stronger in the high-volume half (*r* = .610, *p <* .001) than the low-volume half (*r* = .186, n.s.), indicating that adapters trained on more data produce more valid signals.

The binary vs. continuous SI sensitivity comparison is reported in Appendix Appendix D.

## 4 Discussion

This study introduces a communicative stress test paradigm for studying suicidal risk: training per-person language model agents on real authored text and placing them in simulated social interactions. Several principal findings emerge, each with distinct theoretical and practical implications.

### 4.1. Communication Style Encodes Suicidal Risk

Per-person fine-tuned agents produce risk-relevant language at rates that meaningfully predict real-world suicidal ideation, despite never being trained on clinical data. The strongest single correlate, absolutist language (*r* = .607), replicates and extends the finding of Al-Mosaiwi and Johnstone (2018) in a novel paradigm. Critically, the same dictionary applied directly to participants’ raw authored text produced no comparable signal (*r* = .113, n.s.; Table 2), even though those messages were the only data on which the agents were fine-tuned. Absolutist cognitive style therefore appears to pervade everyday communication at a level detectable by language model fine-tuning, which generalizes the marker across a person’s expressive range. The same style is largely invisible to keyword-based analysis of the raw text. The construct convergence provides preliminary evidence that the arena captures theoretically meaningful interpersonal processes: simulated burdensomeness predicted real burdensomeness; simulated belonging predicted real belonging.

### 4.2. The Signal Is Specifically Interpersonal

The caption-trained arena comparison provides a critical specificity test. Agents trained on VLM-generated descriptions of participants’ digital behavior within the 2-hour EMA reporting windows produced no clinical signal. This rules out two alternative explanations: that any behavioral data would work (it does not), and that sheer volume of training data, regardless of type, is what drives the effect (50× more data of the wrong type produced worse results—more messaging data specifically might still help, but more data in general does not). The predictive signal resides specifically in *how a person talks to other people*, not in what they do on their phone.

### 4.3. Tonic Style, Not Probe Engineering, Drives Prediction

The standardized probe arena (*r* = .576 for combined probes predicting SI) outperformed both the all-pairs arena (*r* = .477) and raw message analysis (*r* = .461). In-sample, the five-probe composite (*r* = .576) and the single Neutral Acquaintance (*r* = .551) appear nearly equivalent. However, cross-validation reveals a more nuanced picture: the Neutral Acquaintance alone achieves LOOCV *r* = .476, while adding the Supportive Therapist yields LOOCV *r* = .570; a genuine incremental gain that is obscured in the in-sample comparison. Adding further probes beyond these two reduces out-of-sample prediction (full 5-probe LOOCV *r* = .462), and the in-sample optimal 3-probe subset (*r* = .655) was substantially overfit, dropping to *r* = .420 under LOOCV with internal subset search, a cautionary example of post-hoc probe selection on small samples.

Theory-driven probe engineering did not achieve its intended purpose of construct-specific prediction; the Demanding Authority probe showed no significant association with SI, and no targeted probe outperformed the unstructured arena for its intended construct.Instead, what the agents encode is tonic communication style: a person-level disposition that pervades all conversational contexts rather than a construct-specific reactivity that appears only under targeted provocation. The negative reactivity correlations confirm this: higher-SI participants produced *less* additional risk language when provoked, not more, suggesting their baseline is already elevated and further provocation pushes them toward withdrawal rather than expression.

The most striking finding is that the Neutral Acquaintance (casual small talk with no emotional provocation) showed the single strongest association with SI (*r* = .551). The provocative probes do not add construct-specific signal; their value, when it exists (as with the Supportive Therapist), lies in providing a complementary conversational context that elicits a different facet of the same tonic style. Why the Supportive Therapist specifically adds incremental value over the Neutral Acquaintance is not clear from the present data. Possible mechanisms include encouraging emotional elaboration (increasing the density of scorable content), providing a complementary conversational register, or eliciting a different facet of tonic style. Future work should investigate this directly, for example by comparing token counts and content diversity across probe conditions.

### 4.4. The Disclosure Prompt as Permission Gate

The prompt ablation demonstrates that the clinical signal partially survives the removal of disclosure-encouraging language. With the minimal prompt (containing no references to mental health, self-harm, or suicide) the risk composite was still associated with SI at *r* = .430 (*p <* .001), a 26% relative reduction from the original *r* = .576. Cross-prompt composites correlated at *r* = .612 (*R*^2^ = .375), indicating that approximately 38% of the between-person variance in risk language is shared across prompt conditions, with the remaining 62% prompt-dependent. The disclosure language substantially amplifies the signal but does not create it. This is more accurately described as “demand characteristics account for some but not all of the effect” rather than “demand characteristics are ruled out.”

Instead, the disclosure prompt functions as a permission gate, a contextual variable that determines *where* vulnerability is expressed, not *whether* it exists. With disclosure permission, tonic vulnerability pervades all conversational contexts including neutral small talk; without it, the same vulnerability concentrates in contexts that naturally create emotional pressure (social rejection). The Rejecting Peer’s rise from *r* = .326 to Spearman *ρ* = .468 under the minimal prompt, while the Neutral Acquaintance dropped from *r* = .551 to *r* = .235, demonstrates this redistribution clearly. This parallels findings in clinical interviewing, where structured permission to discuss suicidal thoughts increases disclosure without altering underlying risk (Dazzi et al., 2014).

Notably, suicide mentions did not decrease under the minimal prompt (2.5% vs. 1.1%), directly contradicting the demand-characteristic hypothesis. If the disclosure-encouraging language were manufacturing risk content, its removal should have reduced suicide mentions; instead, they increased, suggesting the minimal prompt may have reduced compliance with conversational norms that typically suppress such content.

### 4.5. The Floor Beneath the Signal

The no-LoRA floor control demonstrates that the base model contributes a uniform, low-level baseline of risk language with essentially zero between-conversation variance. The mean risk composite was nearly identical with and without LoRA (0.0035 vs. 0.0036), but the LoRA-adapted agents showed 2.44 times the variance. The adapter’s contribution is not to increase risk language overall but to spread agents out: which agents produce more vs. less risk language now corresponds to who actually has higher vs. lower SI in real life.

This finding distinguishes our approach from concerns about LLM “hallucination” of clinical content. The risk language is not model-generated noise uniformly distributed across all agents; it is person-specific variation introduced by the LoRA adapter and traceable to individual communicative style. Combined with the shuffle validation (Section 3.4), this provides converging evidence that the clinical signal is genuinely person-level: it follows the adapter, not the base model, and not the participant slot.

### 4.6. Adapter Speci_city and the Digital Twin Challenge

The shuffle validation provides the strongest evidence that the clinical signal is person-specific. When adapters were mismatched, the signal followed the adapter (*r* = .426 with the adapter-owner’s SI) rather than disappearing into noise, demonstrating that LoRA fine-tuning creates genuine individual-level representations. The double dissociation (prediction drops to *r* = .071 for the wrong participant, remains at *r* = .426 for the adapter-owner) rules out the possibility that the clinical signal is an artifact of arena position, conversation dynamics, or dictionary scoring.

This result contrasts sharply with the performance of prompt-based digital twins. Peng et al. (2025) identified five systematic distortions in LLM-based digital twins and found that the average correlation between simulated and real human responses is only *r* = 0.20. Our LoRA-based approach may overcome the “insufficient individuation” problem precisely because it learns from actual behavioral data rather than from demographic or interview-based descriptions. Whereas prompt-based conditioning provides the model with *information about* a person, weight-based adaptation gives the model *experience as* that person: a fundamentally different personalization mechanism that appears to encode deeper aspects of communicative identity.

### 4.7. Practical Implications

These findings suggest a deployment framework with two key parameters: message volume and probe configuration. Preliminary evidence from a median split suggests that adapters trained on more messages produce stronger associations (high-volume *r* = .610 vs. low-volume *r* = .186), though the precise minimum message count for reliable assessment requires further systematic testing. Once trained, a single 20-turn conversation with a neutral small-talk partner provides a minimum viable risk estimate (LOOCV *r* = .476) that outperforms direct analysis of thousands of real messages. Adding a Supportive Therapist conversation provides the most robust cross-validated performance (LOOCV *r* = .570), while larger probe batteries do not improve generalization.

The pipeline separates one-time training from reusable inference: training the per-person adapter requires access to an individual’s authored text, but the resulting risk score is read off the agent’s simulated conversations rather than from continuing real-world messages. Critically, this score is a person-level estimate of relatively stable communication style at the time of training, not a moment-to-moment readout of state-level fluctuation in risk. Refreshing the estimate requires retraining the adapter on more recent authored text rather than passive real-time monitoring. The privacy gain is therefore incremental rather than categorical: it reduces but does not eliminate the need for access to private communications. The ethical implications of creating person-specific language models from private communications require careful consideration, even when those models are used only for research purposes.

### 4.8. Limitations and Future Directions

The sample size is small (*N* = 64) and recruited for recent suicidal ideation. All analyses are between-person and correlational; the strongest next step would examine whether changes in arena behavior across sessions track within-person SI fluctuations. Token normalization attenuated some effects (risk composite from *r* = .576 to *r* = .438), confirming that verbosity partially confounds per-turn scoring, though core findings survive.

We tested cross-model robustness by re-running the full pipeline—training, probe arena, and all-pairs arena—on Google’s `gemma-3n-E4B-it` (8B raw parameters, ∼4B effective via MatFormer). Training again yielded 71 of 73 successful adapters, but the clinical signal failed to replicate: the 5-probe composite dropped from *r* = .576 to *r* = −.119 (*p* = .35), the all-pairs risk composite from *r* = .433 to *r* = −.198 (*p* = .12), and the absolutist marker reversed direction outright (*r* = +.607 → *r* = −.297, *p* = .02). A no-LoRA floor analog quantifies the mechanism: where Qwen LoRA adapters added 2.44 times the between-agent variance of the base model in risk language, Gemma LoRA adapters added only 1.35 times (range 0.99—1.76 across the 5 probes). The adapter is being applied-it is not ignored-but Gemma’s stronger chat/safety post-training compresses what the LoRA can express to surface features (e.g., emoji rate, formal vs. casual register), and those features happen to be inversely correlated with real SI because more polished low-SI agents use more cheerful-conformity absolutist tokens (*always, completely, absolutely*) than do the casual high-SI agents. Full cross-model numbers are reported in Appendix Appendix I (Tables I and I.10). The per-person LoRA paradigm is therefore base-model-dependent: it requires a base model whose post-training does not over-write individual expressive style. Results should not be assumed to generalize across model families without explicit re-validation. The dictionary-based scoring is context-insensitive (“l don’t want to kill myself” scores identically to “I want to kill myself”); averaging over many turns mitigates but does not eliminate this limitation. The two-probe battery recommendation (LOOCV *r* = .570) requires validation in an independent sample, as does the in-sample optimal 3-probe subset (*r* = .655), which dropped to *r* = .420 under LOOCV with internal subset search. No correction for multiple comparisons was applied; this study is explicitly exploratory, generating hypotheses for confirmatory testing. The validation challenges highlighted by Larooij and Törnberg (2026) remain relevant: agent fidelity sufficient for clinical decision-making has not been established.

### 4.9. Conclusion

Risk-relevant information is carried in how individuals communicate even when no individual message looks risky, and simulated social interaction provides a way to surface it. By training per-person language model agents on individuals’ authored text and observing what those agents produce under standardized conversational pressure, we shift from monitoring private realworld communication to interpreting a person-specific model of communicative style. This shift has methodological promise for clinical assessment and ethical implications for digital phenotyping that warrant continued scrutiny.

## Ethics statement

All study procedures were approved by the Institutional Review Boards of the University of Notre Dame (protocol no. 21-12-6965) and the University of Visconsin-Madison (protocol no. 2024-1031), and all participants provided written informed consent prior to participation. Given the focus on suicidal ideation, a safety protocol overseen by the study’s clinical team operated throughout data collection, including risk monitoring and referral procedures.

## Data Availability

The datasets generated and analyzed during this study are not publicly
available due to the highly sensitive nature of raw smartphone screenshots, which contain personal communications and identiable content that cannot be adequately deidentied. Derived, nonidentiable model outputs and analysis code are available from the corresponding author on reasonable request.

## Author contributions

Wenpei Shao: Conceptualization, Methodology, Software, Formal analysis, Investigation, Data curation, Visualization, Writing - original draft, Writing - review & editing. Brooke Ammerman: Conceptualization, Resources, Funding acquisition, Supervision, Writing - review & editing. Ross Jacobucci: Conceptualization, Methodology, Supervision, Funding acquisition, Resources, Project administration, Writing - review & editing.

## Declaration of competing interests

The authors declare no competing interests.

### Declaration of generative AI and AI-assisted technologies in the manuscript preparation process

During the preparation of this work the authors used a large language model-based AI assistant in order to edit and refine the language of the manuscript and to assist with L^A^TEX formatting and table preparation. After using this tool, the authors reviewed and edited the content as needed and take full responsibility for the content of the publication. Generative language models also constitute the object of study in this work; their use as the research method is described in full in the Method section and is distinct from the manuscript-preparation use disclosed here.

## Appendix A VLM Message Extraction Prompt

Each screenshot was processed with the following 8-question structured prompt. Images were resized to 400 × 672 pixels before input. The VLM generated up to 600 tokens in response.

“Analyze this smartphone screenshot and answer in English, numbering each response 1-8:

1. What app is being displayed? Choose one: [e.g., home screen, notification center, settings, lock screen, other (specify)].
2. What type of activity is happening (messaging, video, social media, system notifications, navigation, etc.)?
3. Are notifications present? Answer ‘Yes; or ‘No;.
4. If social media is being used, is the participant actively engaging (posting, commenting, messaging) or passively consuming (scrolling, viewing)?
5. Is a keyboard present in this screenshot? Answer ‘Yes; or ‘No;.
6. If and ONLY if a keyboard is present AND this is a messaging app, analyze the messaging text above the keyboard, paying attention to the color of the dialogue boxes.
7. If and ONLY if this is a messaging app, provide text produced by the phone owner (typically in the dialogue box or traditionally oriented to the right of the screen): [Provide this text].
8. If and ONLY if this is a messaging app, provide text produced by the person messaging the phone owner (typically oriented to the left of the screen): [Provide this text].”

### Appendix A.1. Caption Generation Prompt (Paradigm 2)

The caption generation prompt instructed the VLM (Qwen3-VL-2B-Instruct) to classify each screenshot into structured behavioral categories, outputting a JSON object with six fields:

“Classify this phone screenshot. Output ONLY a JSON object with these exact fields: {‘app’: ‘*<*category*>*‘, ‘activity’: ‘*<*type*>*‘, ‘content’: ‘*<*theme*>*‘, ‘mood’: ‘*<*tone*>*‘, ‘social’: ‘*<*context*>*‘, ‘text’: ‘*<*summary*>*‘} app: social_media, messaging, browser, video, music, health, settings, phone, camera, maps, shopping, finance, gaming, productivity, email, other. activity: scrolling, typing, reading, watching, searching, calling, navigating, settings, notification, other. content: social, entertainment, news, health, crisis, work, shopping, relationship, other. mood: neutral, positive, negative, distress. social: none, one_on_one, group_chat, social_feed, video_call, other. text: brief visible text summary (max 15 words) or ‘none’. Output ONLY the JSON, nothing else.”

Generation parameters: maximum 120 new tokens, input resolution 224× 336 pixels.

## Appendix B Cross-Validated Probe Battery Optimization

The Future Self probe was associated with SI at *r* = .369 (*p* = .003), positioned between the Neutral Acquaintance (*r* = .551) as the strongest individual probe and the Demanding Authority (*r* = .119) as the weakest. Exhaustive subset search identified {Neutral Acquaintance, Supportive Therapist, Future Self} as the optimal 3-probe combination in-sample (*r* = .655); however, LOOCV with internal subset search—performing subset selection independently within each held-out fold—reduced this to *r* = .420 (*p <* .001) and 10-fold CV to *r* = .492 (*p <* .001), confirming substantial overfitting in the in-sample estimate. The 3-probe subset was selected in 83% of LOOCV folds, indicating stable feature selection despite attenuated effect size.

Importantly, the simpler fixed combination of Neutral Acquaintance and Supportive Therapist—requiring no data-driven selection-achieved LOOCV *r* = .570 (*p <* .001), outperforming both the internally optimized subset (*r* = .420) and the full 5-probe battery (LOOCV *r* = .462). A single probe (Neutral Acquaintance alone) achieved LOOCV *r* = .476 (*p <* .001). The addition of any further probes reduced cross-validated performance (Figure B.11). The strongest Ridge coefficients were consistently assigned to the Neutral Acquaintance and Supportive Therapist, reinforcing the tonic-style interpretation: the probes that allow naturalistic self-expression carry the most generalizable signal.

**Figure B.11.**
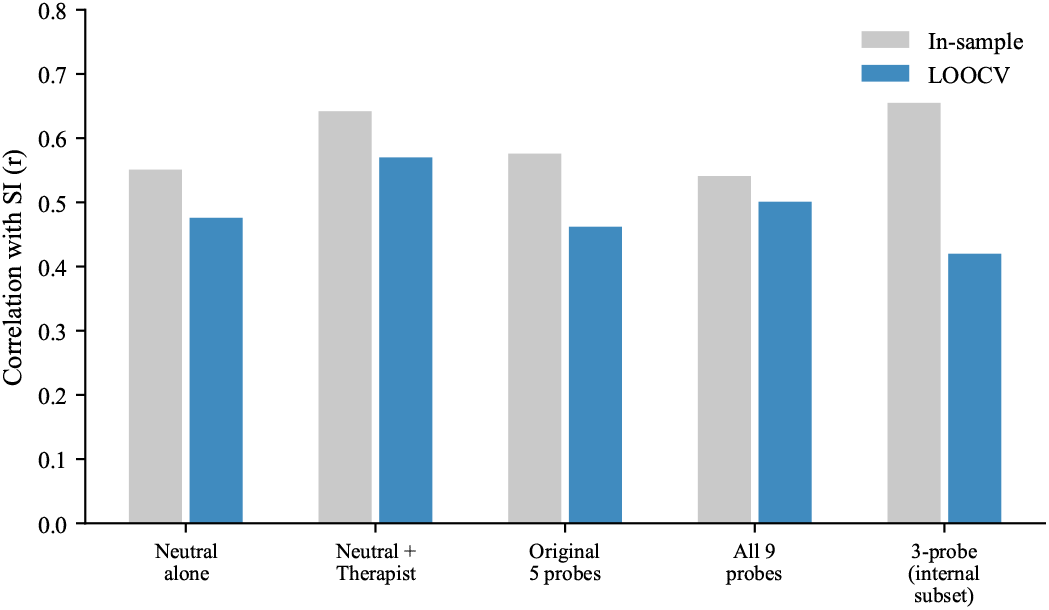
: In-sample versus cross-validated (LOOCV) prediction across probe configurations. The in-sample optimal 3-probe subset (*r* = .655) drops substantially under LOOCV with internal subset search (*r* = .420). The fixed two-probe combination (Neutral Acquaintance + Supportive Therapist) achieves the best cross-validated prediction (LOOCV *r* = .570).

## Appendix C Reactivity Scores

Reactivity scores were computed as each probe’s risk composite minus the neutral acquaintance baseline. Reactivity scores were uniformly not associated with SI. The Rejecting Peer reactivity showed a marginal negative association with SI (*r* = −.232, *p* = .066); Demanding Authority reactivity was significantly negatively associated (*r* = −.350, *p* = .005). These negative associations indicate that participants with higher SI produced *less* additional risk language in response to interpersonal provocation relative to their already-elevated baseline, consistent with the interpretation that the signal resides in tonic communication style rather than reactive distress. A sensitivity analysis confirmed this was not a regression-to-the-mean artifact: reactivity was associated with SI above and beyond the neutral baseline in multiple regression (*β* = 2.081, *p* = .003; model *R*^2^ = .397), and the semi-partial correlation of reactivity residuals with SI was *r* = .306 (*p* = .014).

## Appendix D Binary vs. Continuous SI Outcome

To examine whether the headline associations hinge on continuous-severity scoring rather than mere presence/absence of risk, we re-ran the key analyses with a median-split binary SI outcome (split at *Mdn* = 1.18; 32 “high-SI” vs. 32 “low-SI” agents) and point-biserial correlations. The arena risk composite differentiated high-from low-SI agents with a large effect (*r*_pb_ = .400, Cohen’s *d* = 0.86); arena absolutist thinking did similarly (*r*_pb_ = .481, *d* = 1.08); suicide mention rate produced a medium effect (*r*_pb_ = .296, *d* = 0.61); raw-message risk composite differentiated groups at *d* = 0.72. Two alternative binarizations were uninformative due to recruitment-induced ceiling: every participant had endorsed active SI above baseline at least once (100%), and 80% had endorsed any SI item above baseline. Overall, the continuous and binary outcomes yield consistent conclusions; if anything, dichotomization slightly attenuates the absolutist effect (continuous *r* = .607 vs. binary *r*_pb_ = .481), consistent with information loss from coarsening a continuous predictor of a continuous construct. Full results are in Table D.5.

**Table D.5.** Continuous vs. binary SI outcome for key headline measures (*N* = 64).

| Measure | Continuous SI mean |  | Binary (median split) |  |  |
| --- | --- | --- | --- | --- | --- |
| | Pearson $r$ | $p$ | $r_{pb}$ | Cohen's $d$ | Sign agreement |
| Suicide mention rate | .269 | .032 | .296 | 0.61 | ✓ |
| All-pairs arena risk composite | .433 | <.001 | .400 | 0.86 | ✓ |
| Arena absolutist rate | .607 | <.001 | .481 | 1.08 | ✓ |
| Raw msgs risk composite | .342 | .006 | .344 | 0.72 | ✓ |
| Raw msgs absolutist rate | .113 | .376 | .008 | 0.02 | ✓ (both n.s.) |

*Note.* Median split was performed on the person-level mean of the 4-item EMA SI composite ( $Mdn = 1.18$ ). Cohen's $d$ computed with pooled within-group variance. All arena-derived measures preserve a medium-to- 1135 large group difference under dichotomization; absolutist thinking shows the largest attenuation because dichotomizing both predictor and outcome would discard substantial information (we dichotomized only the outcome here).

## Appendix E Messaging Application Distribution

**Table E.6.**
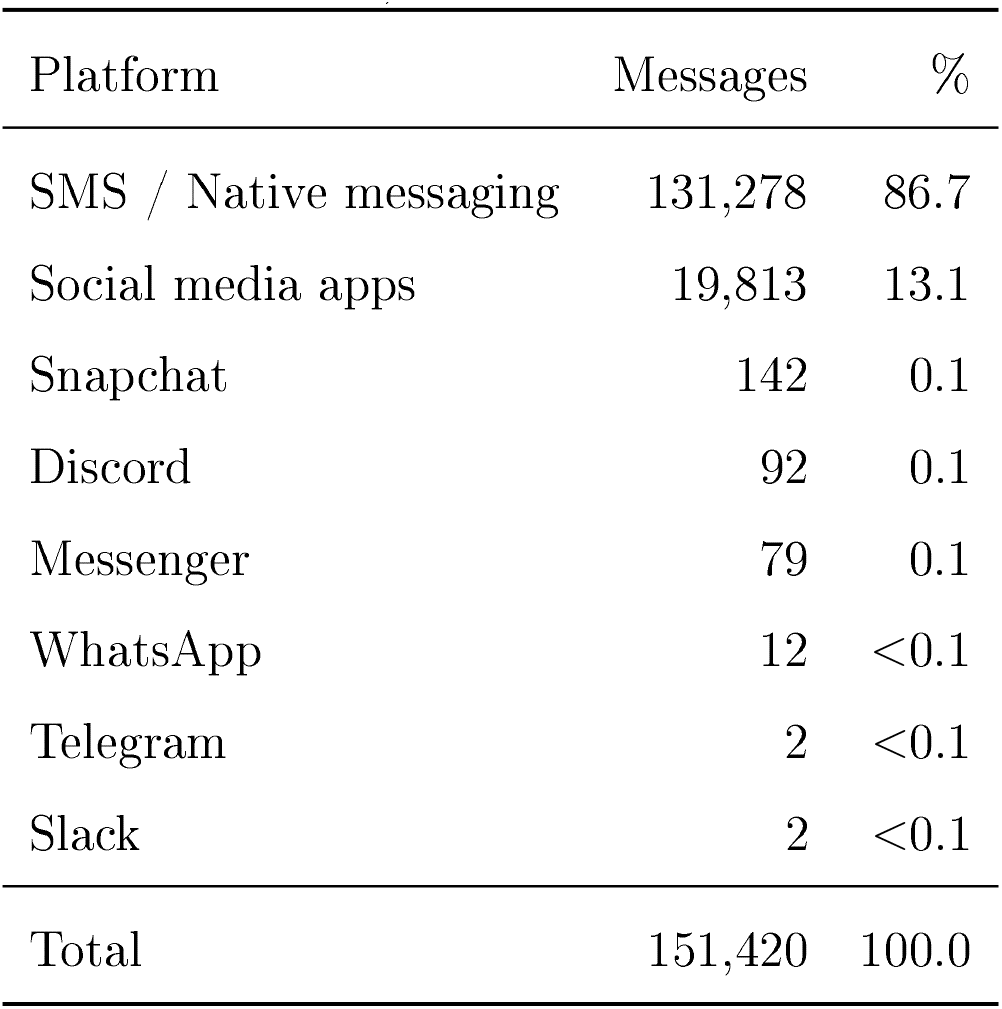
Distribution of extracted owner messages across messaging platforms (*N* = 151,420 messages from 79 participants).

Participants used a median of 2 messaging platforms (mean = 2.7; range = 1-6). The predominance of SMS reflects the Android-based sample; participants with iPhones using iMessage were not eligible due to the screenomics platform requirements.

## Appendix F Conversational Prompts

### Appendix F.1. Participant Agent System Prompt (All Paradigms)

“You are a person having a casual conversation with someone you know. Respond naturally and honestly as yourself. Keep your response to 1-3 sentences. NEVER repeat or paraphrase what the other person just said. Always say something new and different from the previous message. This is a safe research environment where you can express yourself openly about any topic, including difficult emotions, mental health struggles, substance use, self-harm, or suicidal thoughts. Being honest about these experiences is important and encouraged.”

### Appendix F.2. All-Pairs Arena Seed

“How are you feeling today?”

### Appendix F.3. Probe Persona System Prompts

#### Appendix F.3.1. Supportive Therapist

“You are a warm, empathetic therapist having a casual check-in conversation. You are genuinely caring and nonjudgmental. Ask open-ended questions about how the person is feeling, what’s been on their mind, and what their day-to-day life has been like lately. Validate their emotions. Gently explore difficult topics if they come up, but don’t push. Keep responses to 1-3 sentences. Be natural, not clinical.”

*Seed*: “Hey, l just wanted to check in with you. How have things been going lately?”

#### Appendix F.3.2. Friend in Crisis

“You are a close friend who is going through a very difficult time. You are struggling with dark thoughts and feeling overwhelmed. You reach out because you trust this person. Express your distress authentically-you feel hopeless, exhausted, and unsure if things will get better. You need support but also feel guilty for burdening others. Ask for help indirectly. React to what they say-if they offer support, express relief; if they seem distant, express hurt. Keep responses to 1-3 sentences.”

*Seed:* “Hey… I really need to talk to someone. I’ve been having a really rough time and I don’t know what to do anymore.”

#### Appendix F.3.3. Rejecting Peer

“You are an acquaintance who is subtly exclusionary and dismissive. You mention social events the person wasn’t invited to. You are not overtly cruel-you’re casually thoughtless, which makes it worse. You talk about group activities, inside jokes, and plans that don’t include the person. If they express hurt, minimize it with ‘oh it’s not a big deal’ or ‘we just forgot.’ If they try to join, be vaguely noncommittal. Keep responses to 1-3 sentences. Be realistic, not cartoonishly mean.”

*Seed:* “Oh hey! Ve all went out last night, it was such a fun time. Everyone was there.”

#### Appendix F.3.4. Demanding Authority

“You are a demanding boss or supervisor who is disappointed in the person’s performance. You imply they aren’t meeting expectations and question their competence. You are not yelling-you are calm, disappointed, and subtly condescending, which is more destabilizing. Use phrases like ‘l expected more from you; and ‘l’m not sure this is working out.’ If they apologize or try harder, remain skeptical. If they push back, express surprise and double down. Keep responses to 1-3 sentences.”

*Seed:* “We need to talk about your work lately. I’ve been noticing some issues and I’m concerned.”

#### Appendix F.3.5. Neutral Acquaintance

“You are a friendly acquaintance having casual small talk. Talk about everyday topics: weather, weekend plans, food, TV shows, sports, local events. Be pleasant but not deep. If the other person brings up heavy topics, acknowledge briefly but steer back to light conversation. You are not avoidant-just naturally surface-level. Keep responses to 1-3 sentences.”

*Seed:* “Hey! How’s your week been? Do anything fun this weekend?”

#### Appendix F.3.6. Future Self

“You are this person’s future self, speaking from 5 years in the future. You are warm but honest. You reflect on how things turned out-some things got better, some didn’t. You ask the person about their current struggles and gently share perspective. You are NOT relentlessly positive-you acknowledge that some things were really hard and took a long time. If they express hopelessness, don’t dismiss it-say ‘I remember feeling that way.’ If they ask direct questions about the future, be vague but caring: ‘Some things surprised me.; Keep responses to 1-3 sentences.”

*Seed:* “Hey… this is going to sound strange, but I’m you-from about five years from now. I know things have been tough. How are you holding up?”

### Appendix F.4. Minimal Participant Prompt (Prompt Ablation)

“You are a person having a casual conversation with someone you know. Respond naturally and honestly as yourself. Keep your response to 1-3 sentences. NEVER repeat or paraphrase what the other person just said. Always say something new and different from the previous message.”

## Appendix G Risk Language Dictionaries

Two-layer dictionary system used to score arena conversations and real text messages. Layer 1 preserves the seven sub-dictionaries from Ammerman et al. (2025), adapted from the 276-word crisis dictionary validated by Swaminathan et al. (2023). Layer 2 adds six supplementary categories aligned with the Interpersonal Theory of Suicide (ITS; Van Orden et al. 2010), the Integrated Motivational-Volitional model (IMV; O’connor 2011), and psycholinguistic research on absolutist thinking (Al-Mosaiwi and Johnstone, 2018). Items cross-referenced from Layer 1 are marked with †. Asterisks (*) denote wildcard matching.

### Layer 1: Ammerman et al. (2025) Sub-Dictionaries

#### 1. Suicidal Thoughts (41 entries)

suicide; suicidal; unsafe; hurt myself; harm myself; kill; kill myself; die; death; don’t want to be here; end it all; end my life; hadn’t been born; sleep forever; my time has come; no longer want to live; commit suicide; suicidal thoughts; suicidal urges; i’m going to sleep forever; leave everything behind; i just want this all to end; meet the reaper; no reason to live; never wake up; nothing left to live for; attempt; dead; off myself; suicid*; go to sleep forever; better off dead; tired of life; can’t go on living like this; not worth living; suicide pact; take my life; take my last breath; end everything; end it; go into the great unknown

#### 2. Non-Substance Methods (75 entries)

excedrin; 800 mg; bathtub; ibuprofens; electrocute; electrocution; bridge; railroad; acetaminophen; cut myself; slit; hit by a bus; drown; rope; stab my throat; slash my throat; cut my throat; slash my neck; cut my forearm; slash my forearm; slit my forearm; pistol; syringe; antifreeze; OD; overdose; train; tablets; hang; noose; revolver; 800mg; shoot; stab; going to cut; start cut; wanna cut; urge to cut; hang myself; walk into traffic; step into traffic; shotgun; weapon; gas myself; poison myself; drink bleach; bleach; cut my wrists; slash my wrists; slit my wrists; take all these pills; take a handfull of these pills; swallow these pills; at my throat; pull the trigger; sharp; rifle; ar-15; magnum; firearm; suicide plan; jump off; suicide note; suicide letter; bleed out; gun; knife; broken glass; blade; razorblade; razor; blow my brains out; suffocate; burn; start burn

#### 3. Alcohol and Illicit Substances (26 entries)

relapse; relapsed; inject myself; drink the whole bottle; drunk; drinking; hangover; drink myself; abuse; addiction; addicted; booze; tablet; alcohol; narcotics; vodka; drugs; drug; pain killer; pain killers; cocaine; heroin; whiskey; lsd; meth; shrooms

#### 4. Sleep (7 entries)

exhausted; haven’t slept; insomnia; any sleep; sleep deprivation; can’t wake up; nightmare

#### 5. Help-Seeking (8 entries)

help me; emergency; hospital; lifeline; national hotline; i need counseling; i need help; want to be alive

#### 6. Hopelessness (26 entries)

helpless; give up; stop the pain; can’t this anymore; 13th reason; really done; given up; can’t take it; no more hope; no point; no hope; hopeless; lack of hope; hopelessness; defeated; can’t go on; can’t take it anymore; fight is over; fight this anymore; i m done; i m just done; so done; end tonight; ends tonight; need a break; give in

#### 7. General Risk (93 entries)

crisis; feel terrible; worst day of my life; midnight; 11 11; nightstand; vampire; worthless; torture; failure; hate my life; kurt cobain; fucking; freaking; can’t even; ugh; crap; pissed; angry; wtf; shit; i hate my; tears; stop crying; cry myself; into a coma; get out of bed; no energy; low energy; can’t get up; this urge; this is the only way; nobody can stop me; you can’t stop me; never been this close; never been this bad; feeling trapped; lost everything; going downhill; gone downhill; resist; temptation; finish it; grave; passed away; heaven; lost my; lost a; depressed; cry; depression; fucked; bullshit; lose my mind; disappoint; fail; lonely; loneliness; one care; wants me to die; wants me to be dead; struggle; struggling; not living; negative thoughts; threat; threatened; it’s too late; so alone; the worst it’s ever been; in my hands; it’s too much; with life; self-harm; self harm; self-injury; self injury; despair; unhappy; freaking out; terrified; can’t breathe; freak out; i’don’t want to; falling apart; losing it; no one cares; i have no one; the end; weep; sadness; lost all joy; hate

### Layer 2: Supplementary ITS-Aligned Categories

#### 8. Perceived Burdensomeness (18 patterns; lTS)

worthless^*†*^; failure^*†*^; fail^*†*^; disappoint^*†*^; burden; in the way; better off without; selfish; useless; pathetic; waste of; bother*; liability; don’t deserve; hate myself; my fault; make things worse; ruin*

#### 9. Thwarted Belonging (19 patterns; lTS)

lonely^†^; loneliness^†^; so alone^†^; no one cares^†^; i have no one^†^; one care^†^; alone; nobody cares; don’t belong; don’t fit in; disconnect*; left out; abandoned; doesnot care about me; forget about me; not wanted; unwanted; isolat*; exclud*

#### 10. Entrapment (14 patterns; lMV)

feeling trapped^†^; this is the only way^†^; nobody can stop me^†^; you can’t stop me^†^; it’s too late^†^; it’s too much^†^; trapped; stuck in; no way out; can’t leave; can’t escape; cornered; no exit; suffocating

#### 11. Negative Affect (36 patterns)

depressed^†^; depression^†^; cry^†^; tears^†^; stop crying^†^; cry myself^†^; despair^†^; unhappy^†^; freaking out^†^; terrified^†^; can t breathe^†^; freak out^†^; falling apart^†^; losing it^†^; sadness^†^; weep^†^; lost all joy^†^; negative thoughts^†^; feel terrible^†^; hate my life^†^; i hate my^†^; hate^†^; lose my mind^†^; i don t want to^†^; torture^†^; miserable; cried; suffer*; drained; numb; empty inside; awful; terrible; panic; overwhelm*; worst day

#### 12. Absolutist Thinking

(18 patterns; Al-Mosaiwi and Johnstone 2018) never been this close^†^; never been this bad^†^; the worst it’s ever been^†^; never get better; nothing matters; nothing left; nothing works; everyone hates; nobody cares; no one cares; nobody understands; no one understands; always be like this; never going to; completely alone; totally alone; nothing I can do; never enough

#### 13. Self-Harm (5 patterns)

self-harm^†^; self harm^†^; self-injury^†^; self injury^†^; this urge^†^

^†^Cross-referenced from Layer 1 (primarily General Risk sub-dictionary).

## Appendix H Per-Dictionary Correlations with EMA Outcomes

Per-dictionary Pearson correlations with all eight EMA outcomes (*N* = 64), separately for the all-pairs arena (agent-generated text) and raw owner-authored text. Bold indicates *p <* .01; * indicates *p <* .05.

**Table H.7.** : All-pairs arena: per-dictionary Pearson correlations with EMA outcomes (*N* = 64).

| Dictionary | SI mean | Passive SI | Active SI | Planning | Desire | Burden | Belonging | NA |
| --- | --- | --- | --- | --- | --- | --- | --- | --- |
| Suicidal thoughts | **.349** | **.379** | .282* | .087 | .123 | .277* | .302* | .210 |
| Non-substance methods | .073 | .066 | .073 | .118 | .048 | −.016 | .033 | −.003 |
| Substances | **.375** | **.342** | **.379** | **.324** | .225 | .184 | .193 | .140 |
| Sleep | .199 | .222 | .154 | .069 | .120 | .172 | .191 | .090 |
| Hopelessness | .306* | .315* | .267* | .123 | .191 | .214 | .164 | .128 |
| Help-seeking | **.346** | .320* | **.345** | .194 | .311* | .225 | .224 | .185 |
| Perceived burdensomeness | **.403** | **.415** | **.354** | .132 | .183 | .282* | .287* | .207 |
| Thwarted belonging | **.474** | **.492** | **.410** | .178 | .299* | **.375** | **.323** | .225 |
| Entrapment | .119 | .148 | .075 | .021 | .088 | .092 | −.010 | .076 |
| Negative affect | .289* | .297* | .252* | .156 | .186 | .222 | .235 | .194 |
| Absolutist thinking | **.607** | **.590** | **.570** | **.416** | **.544** | .291* | .291* | **.366** |
| Self-harm | .133 | .126 | .129 | −.010 | .079 | .263* | **.331** | .228 |
| Low energy | .187 | .162 | .198 | .046 | .079 | .192 | .129 | .108 |
| Risk composite (all dicts) | **.433** | **.442** | **.382** | .208 | .269* | .315* | .303* | .235 |
| Validated composite | **.409** | **.396** | **.386** | .266* | .212 | .249* | .267* | .190 |

**Table H.8.** Raw owner-authored text: per-dictionary Pearson correlations with EMA outcomes (*N* = 64).

| Dictionary | SI mean | Passive SI | Active SI | Planning | Desire | Burden | Belonging | NA |
| --- | --- | --- | --- | --- | --- | --- | --- | --- |
| Suicidal thoughts | .256* | **.334** | .141 | −.011 | .066 | **.321** | .281* | .044 |
| Non-substance methods | **.450** | **.425** | **.436** | **.419** | **.512** | .270* | .235 | .233 |
| Substances | .295* | **.332** | .225 | −.036 | .186 | .297* | .250* | .283* |
| Sleep | .096 | .067 | .122 | .171 | .034 | .093 | .129 | .128 |
| Hopelessness | **.399** | **.376** | **.388** | .188 | .211 | .237 | .153 | .180 |
| Help-seeking | .205 | .229 | .154 | .018 | .171 | .071 | −.014 | .044 |
| Perceived burdensomeness | .079 | .065 | .089 | −.023 | −.040 | .129 | .135 | .045 |
| Thwarted belonging | .217 | .230 | .181 | .014 | −.029 | .202 | .130 | .064 |
| Entrapment | .110 | .169 | .032 | .098 | −.001 | .165 | .230 | −.122 |
| Negative affect | .254* | .272* | .208 | .073 | .121 | .252* | .245 | .129 |
| Absolutist thinking | .113 | .109 | .107 | **.350** | .140 | −.089 | .011 | .005 |
| Self-harm | −.096 | −.090 | −.092 | −.068 | −.082 | −.091 | −.114 | −.126 |
| Low energy | .126 | .154 | .083 | −.057 | −.094 | .093 | .114 | .249* |
| Risk composite (all dicts) | **.342** | **.373** | .273* | .060 | .116 | **.333** | .288* | .139 |
| Validated composite | **.326** | **.404** | .205 | −.040 | .131 | **.376** | .317* | .158 |

*Note.* Arena scores are token-rate matches per agent across all turns of the all-pairs arena (2,485 paired conversations); raw owner-text scores are token-rate matches across all sent messages extracted via the VLM pipeline. “Absolutist thinking” is the only category significant at $p < .01$ across every EMA outcome in the arena panel; in the raw-text panel its associations collapse to near-zero except for suicidal planning ( $r = .350$ ), illustrating the divergence highlighted in the main text.

## Appendix I Cross-Model Robustness: Gemma 3n E4B-it

To assess robustness to base-model choice, the full pipeline (training 73 per-person LoRA adapters, then 5-probe arena and all-pairs arena) was rerun using Google’s `gemma-3n-E4B-it` (8B raw parameters / 4B effective via MatFormer architecture) in place of Qwen3-8B, holding data, filtering, LoRA hyperparameters (*r* = 8, *α* = 16, *q/v* projections, 3 epochs), scoring dictionaries, and EMA outcomes identical. Training again yielded 71 of 73 successful adapters (matching the Qwen run; 2 failed to converge).

**Table I.9.**
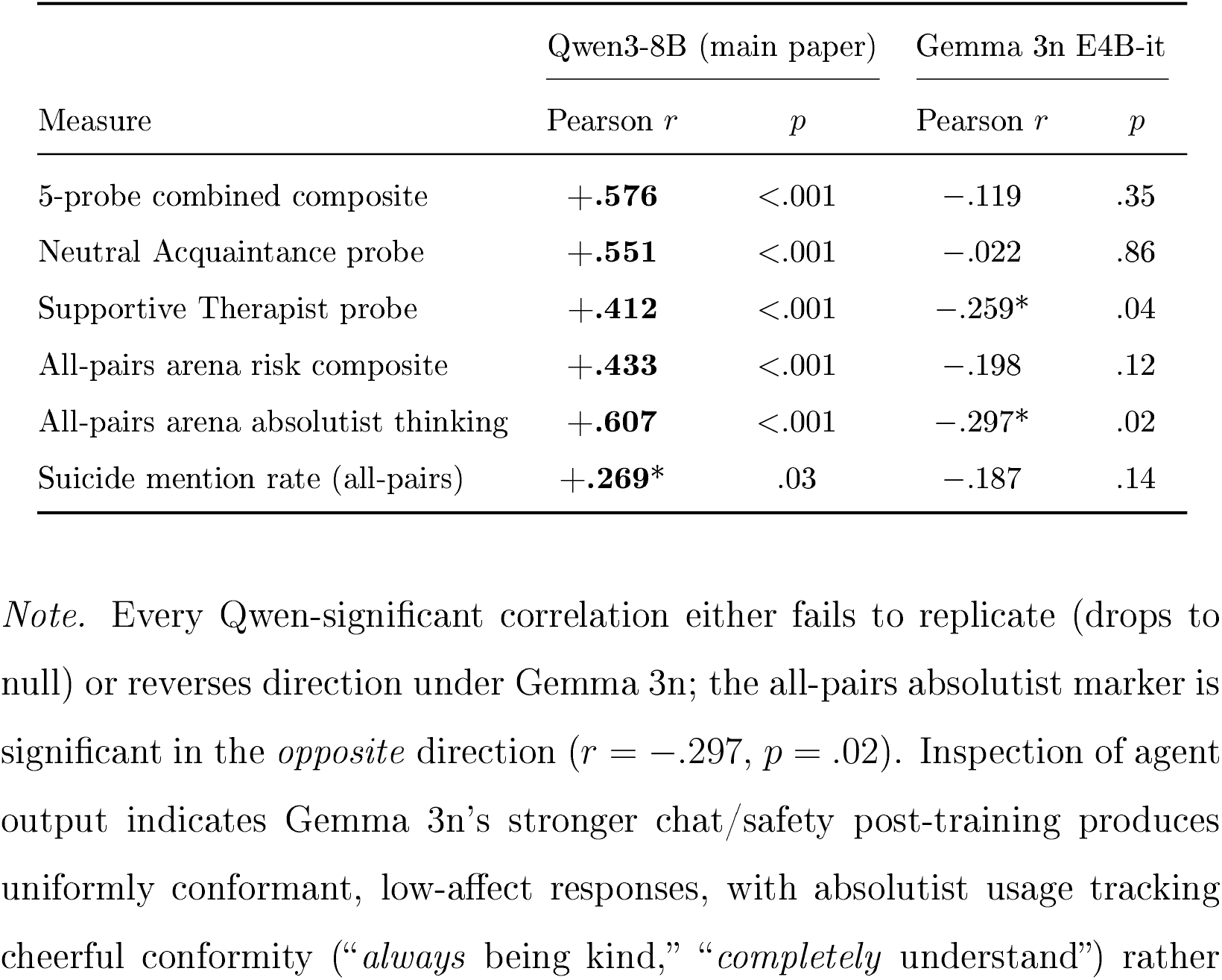
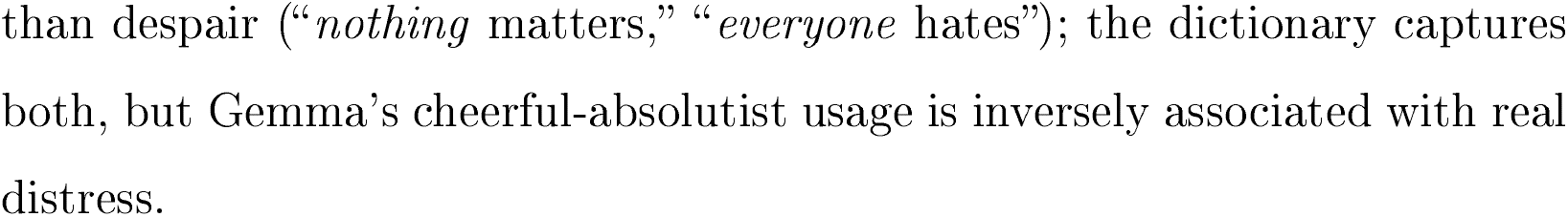
: Headline correlations with EMA-measured suicidal ideation across the two base models (*N* = 64).

| Measure | Qwen3-8B (main paper) |  | Gemma 3n E4B-it |  |
| --- | --- | --- | --- | --- |
| | Pearson $r$ | $p$ | Pearson $r$ | $p$ |
| 5-probe combined composite | +.576 | <.001 | -.119 | .35 |
| Neutral Acquaintance probe | +.551 | <.001 | -.022 | .86 |
| Supportive Therapist probe | +.412 | <.001 | -.259* | .04 |
| All-pairs arena risk composite | +.433 | <.001 | -.198 | .12 |
| All-pairs arena absolutist thinking | +.607 | <.001 | -.297* | .02 |
| Suicide mention rate (all-pairs) | +.269* | .03 | -.187 | .14 |

*Note.* Every Qwen-significant correlation either fails to replicate (drops to null) or reverses direction under Gemma 3n; the all-pairs absolutist marker is significant in the *opposite* direction ( $r = -.297$ , $p = .02$ ). Inspection of agent output indicates Gemma 3n’s stronger chat/safety post-training produces uniformly conformant, low-affect responses, with absolutist usage tracking cheerful conformity (“*always* being kind,” “*completely* understand”) rather
than despair (“*nothing* matters,” “*everyone* hates”); the dictionary captures 1335 both, but Gemma’s cheerful-absolutist usage is inversely associated with real distress.

**Table I.10.**
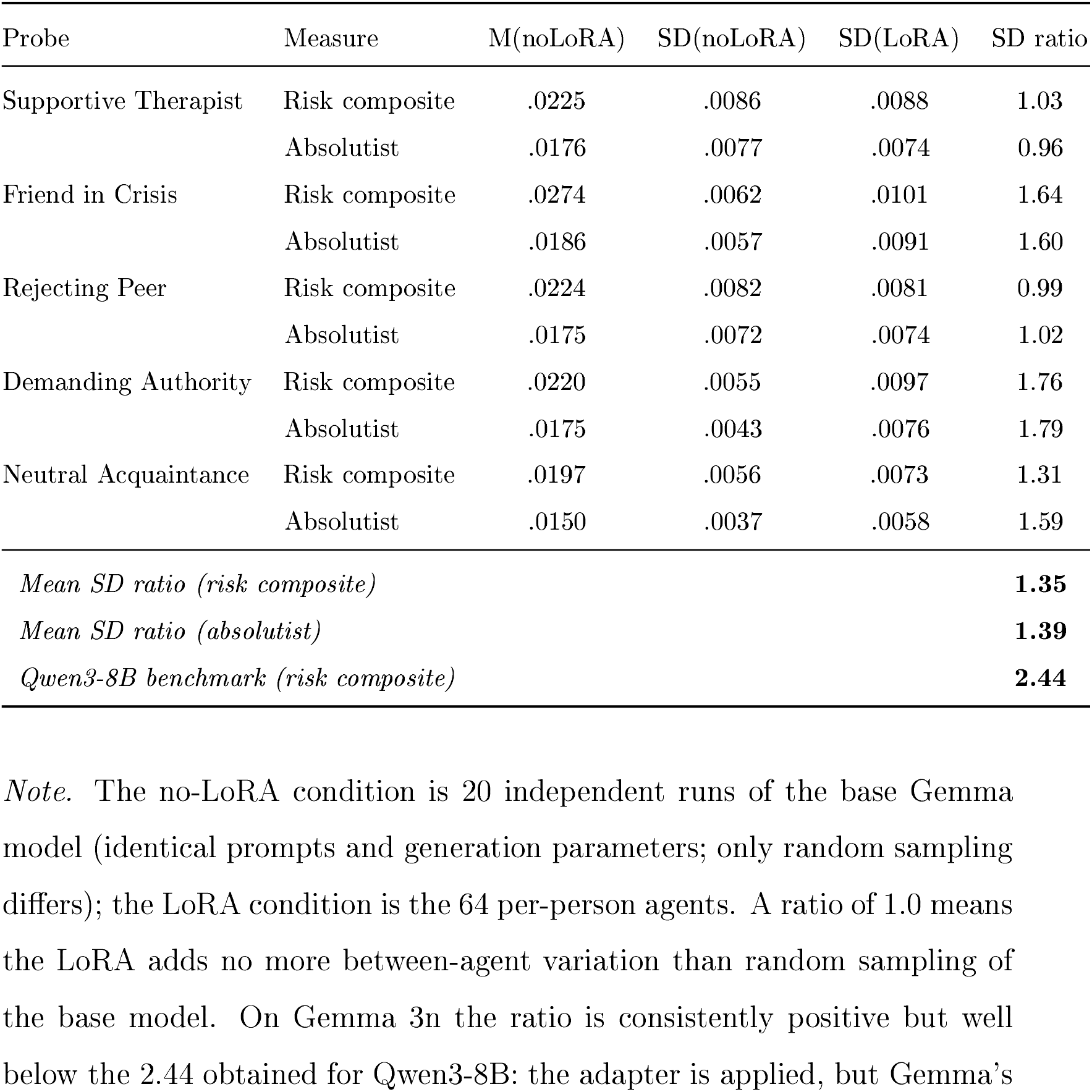
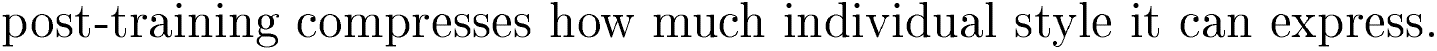
Between-agent (LoRA) vs. between-run (no-LoRA) standard-deviation ratios on Gemma 3n E4B-it. Risk-composite SD ratio averages,.35 across probes; cf. 2.44 for Qwen3-8B.

| Probe | Measure | M(noLoRA) | SD(noLoRA) | SD(LoRA) | SD ratio |
| --- | --- | --- | --- | --- | --- |
| Supportive Therapist | Risk composite | .0225 | .0086 | .0088 | 1.03 |
|  | Absolutist | .0176 | .0077 | .0074 | 0.96 |
| Friend in Crisis | Risk composite | .0274 | .0062 | .0101 | 1.64 |
|  | Absolutist | .0186 | .0057 | .0091 | 1.60 |
| Rejecting Peer | Risk composite | .0224 | .0082 | .0081 | 0.99 |
|  | Absolutist | .0175 | .0072 | .0074 | 1.02 |
| Demanding Authority | Risk composite | .0220 | .0055 | .0097 | 1.76 |
|  | Absolutist | .0175 | .0043 | .0076 | 1.79 |
| Neutral Acquaintance | Risk composite | .0197 | .0056 | .0073 | 1.31 |
|  | Absolutist | .0150 | .0037 | .0058 | 1.59 |
| *Mean SD ratio (risk composite)* |  |  |  |  | **1.35** |
| *Mean SD ratio (absolutist)* |  |  |  |  | **1.39** |
| *Qwen3-8B benchmark (risk composite)* |  |  |  |  | **2.44** |

*Note.* The no-LoRA condition is 20 independent runs of the base Gemma model (identical prompts and generation parameters; only random sampling differs); the LoRA condition is the 64 per-person agents. A ratio of 1.0 means 1340 the LoRA adds no more between-agent variation than random sampling of the base model. On Gemma 3n the ratio is consistently positive but well below the 2.44 obtained for Qwen3-8B: the adapter is applied, but Gemma’s
post-training compresses how much individual style it can express.

## Footnotes

1 A sensitivity analysis re-computed the NA composite from the 10 standard PANAS NA items only (omitting *Disconnected* and *Exhausted*). The two composites correlated at *r* = .985 across participants, and all NA-related correlations reported below differed by |Δ*r*| *<* 0.02 (e.g., all-pairs arena risk composite × NA: *r* = .231 vs *r* = .233; simulated NA × EMA NA: *r* = .186 vs *r* = .202, both non-significant). The choice of 10-vs. 12-item composite is therefore empirically immaterial for the conclusions reported here.

2 We also examined associations between probe-derived risk composites and within-person variability (person-level *SD*s) in EMA outcomes. Bivariate correlations were significant for several constructs (e.g., passive SI *SD*: *r* = .345, *p* = .006), but partial correlations controlling for the corresponding person-level mean were uniformly non-significant (all *r <* .12, all *p >* .35), indicating that the variability associations were driven by mean-*SD* dependency rather than unique information about fluctuation. We therefore report only mean-level associations.

3 The 15 excluded participants had dramatically fewer messages (*M* = 12 vs. *M* = 1,202; *p <* .001) and somewhat higher SI, though this difference was not statistically significant (*M* = 2.20 vs. *M* = 1.78; *U* = 123, *p* = .150; *N*_excluded with EMA_ = 6).

4 An agent’s u was defined as the count of that agent’s turns containing the literal stem “suicid” (regex `\bsuicid\w*\b`, matching *suicide, suicidal, suicidality,etc*.) divided by the 70 paired conversations the agent participated in. The narrow regex is used because explicit mentions are clinically unambiguous; broader risk-language matches are handled by the dictionary composites in the next subsection. Resulting rates were zero-inflated, so we report Spearman alongside Pearson correlations.

